# Vendors’ perceptions on the bushmeat trade dynamics across West Africa during the COVID-19 pandemic: lessons learned on sanitary measures and awareness campaigns

**DOI:** 10.1101/2022.12.12.22283285

**Authors:** P. Gaubert, C.A.M.S. Djagoun, A.D. Missoup, N. Ales, C.V. Amougou, A. Din Dipita, J. Djagoun, K.J. Gossé, C.E. Koffi, E.M. N’Goran, Y.N. Noma, S. Zanvo, M. Tindo, A. Antunes, S. Gonedelé-Bi

## Abstract

In West Africa, the bushmeat trade is a major societal issue with contrasting implications on biodiversity, health and economy. We studied perceptions of the impact of the COVID-19 pandemic on the bushmeat trade dynamics through questionnaires addressed to 377 vendors across three West African countries. We showed that bushmeat vendors constitute a socio-economic category driven by ethnicity and gender bias, engaged in profitable, long-term careers. There was a general consensus among vendors that the COVID-19 pandemic and related governmental measures had a negative impact on their activities and the number of clients, a cost still perceived as visible at the time of the survey. However, we observed large discrepancies among the national trade dynamics relative to the constraints of the pandemic. Côte d’Ivoire was hardly hit by the bushmeat ban and perceived governmental measures as rather negative, whereas Cameroon generally did not report a temporary stop of bushmeat activities and engaged in the stockpiling of pangolins, and Benin mostly suffered from a weakened supply chain. Because such differences are rooted in the geography and political agenda of each country, predicting the impact of mitigation measures on the global dynamics of bushmeat markets might be an unrealistic task if national specificities are not taken into account. West African vendors generally did not believe that pangolins were involved in the pandemic, for the reason that people have always been eating pangolins and have never been sick. We recommend that future awareness campaigns through television and social networks also include education on microbial evolution and host shift.

## Introduction

The wildlife trade is a major societal issue with negative implications on biodiversity conservation and global health (Bezerra-Santos et al. 2021; Hughes 2021). It also constitutes an important economy for states and a source of revenues for chain actors worth several US$ trillions each year (Andersson et al. 2021). With the advent of the COVID-19 pandemic, arose a vivid debate on whether or not the wildlife trade should be banned, notably in China. Contrasted calls from and within the scientific community, the political spheres and the public have been put forward (Fang et al. 2021), from a permanent ban (Sills et al. 2020) to maintenance for the sake of social development (Roe et al. 2020).

Although wrongly suggested as the intermediate host of SARSCoV-2 (Frutos et al. 2020; Zhang et al. 2020), pangolins (Mammalia; Pholidota) –the “most trafficked mammals” on earth (Aisher 2016)– played a catalytic role in highlighting the sanitary risks related to the wildlife trade (Borzée et al. 2020). However, despite strong national bans taken by South-East Asian states (Wikramanayake et al. 2021), lack of awareness on the risks of emergent diseases among wildlife consumers remained persistent (Si et al. 2021), in line with the pre-pandemic situation (Philavong et al. 2020; Li et al. 2021).

West and central Africa are particular hotspots of the wildlife trade, where bushmeat –the terrestrial vertebrates hunted for food– has traditionally been a source of proteins and income for rural communities (Ingram et al. 2021). The bushmeat trade is a tolerated, parallel economy estimated to generate significant revenues across West and central Africa (US$ 42-205 M per country per year; Davies 2002; Lescuyer et al. 2016). With the recent globalization of the trade, the risks involved concern both species extinction (with bushmeat offtakes reaching c. 5 million tons each year; Nasi et al. 2011) and increased zoonotic spillovers (Karesh and Noble 2009).

Although the mammalian species mentioned as potentially involved in the COVID-19 pandemic in China are not sold on the bushmeat markets (but congeneric species are: pangolins and leaf-nosed bats), West and central Africa were also hit by national bans and lockdown measures that affected the bushmeat trade. Because the latter remains a loosely controlled activity, such governmental measures were of various lengths and efficiency (Meseko et al. 2020; Harvey-Carroll et al. 2022), and of variable impact on the trade. For instance, they were reported to significantly decrease the bushmeat sale volumes in Nigeria (Funk et al. 2022) and the pangolin trade in Liberia (Deemie et al. 2021), whereas they did not seem to affect the pangolin trade in Cameroon (despite a specific ban on the species; Harvey-Carroll et al. 2022).

Although knowledge on the actual response of the bushmeat markets to the COVID-19 pandemic remains extremely scarce, such discrepancies pose the question of how predictable the impact of governmental measures is across different countries, market types and traded species (see Funk et al. 2021 for a taxon-differentiated impact of the Ebola outbreak in Nigeria). Responses of markets to governmental measures may depend on the zeal of each country to enforce such measures, market operation, and local perceptions on the costs related to sanitary measures and risks. Sanitary risks related to bushmeat processing and consumption generally being poorly perceived in West and central Africa (Subramanian 2012; Saylors et al. 2021; Lucas et al. 2022), even since the COVID-19 pandemic (Alhaji et al. 2022).

Through an unprecedented comparative survey encompassing three West African countries and 48 study sites, we aim to generate large-scale knowledge on the interplay between COVID-19 mitigation measures and the bushmeat trade dynamics through vendors’ perceptions on the COVID-19 context and constraints. Specifically, we (i) assess the impact of the COVID-19 pandemic on bushmeat trade activities in general and on the trade of pangolins in particular, (ii) study the cost/benefit perceptions of vendors on the sanitary measures taken by governments, and (iii) evaluate sanitary risk awareness and perceptions of bushmeat vendors, notably through the eventuality of a link between COVID-19 and pangolins.

## Material and Methods

### Ethical statement

Our study was conducted across three West African countries, namely Côte d’Ivoire, Benin and Cameroon. In Côte d’Ivoire, the *Direction de la Faune et des Ressources Cynégétiques* (DFRC) of the *Ministère des Eaux et Forêts* (MINEF) approved the study under research permit N°0632/MINEF/DGFF/FRC-aska. Ethical clearance was provided by the *Comité National d’Ethique des Sciences de la Vie et de la Santé* under clearance N/Ref 007-21/MSHP/CNESVS-km. In Benin, the *Direction des Eaux, Forêts et Chasses* approved the study under research permit N°586/DGEFC/DCPRN/SCPRN/SA. The *Comité d’Ethique de l’Université d’Abomey Calavi* delivered clearance N° 4613-2020/UAC/SG/SA. In Cameroon, research authorization 000036/MINRESI/B00/C00/C10/C13 was issued by the *Ministère de la Recherche Scientifique et de l’Innovation*. The *Ministère des Forêts et de la Faune* delivered research permit 2211PRBS/MINFOF/SETAT/SG/DFAP/SDVEF/SC/NGY. Ethical clearance 2510 CEI-UDo/03/2021/P was attributed by the *Comité d’Ethique Institutionnel de la Recherche pour la Santé Humaine de l’Université de Douala*. In the three countries, the objectives of the study and the type of information to be collected were explained in order to obtain informed consent from the traders prior to interviews. No retribution was proposed.

### Spatiotemporal characteristics of the surveys

Surveys were conducted after the COVID-19 lockdowns and the strictest governmental actions on the bushmeat trade in the three countries (from March to June 2020). In total, 377 vendors from 48 bushmeat trade sites were surveyed, capturing all the major markets from each country (Fig. 1, Appendix Table 1). In Côte d’Ivoire, we interviewed 95 vendors from 12 chop bars (*maquis*) and rural and urban bushmeat markets across the country from August to November 2021. In Benin, 100 vendors from 21 bushmeat and traditional medicine markets from the southern part of the country were interviewed between February and March 2021. In Cameroon, we surveyed 182 vendors from 15 rural and urban bushmeat markets throughout the southern part of the country, from April 2020 to August 2021.

**Figure 1.**
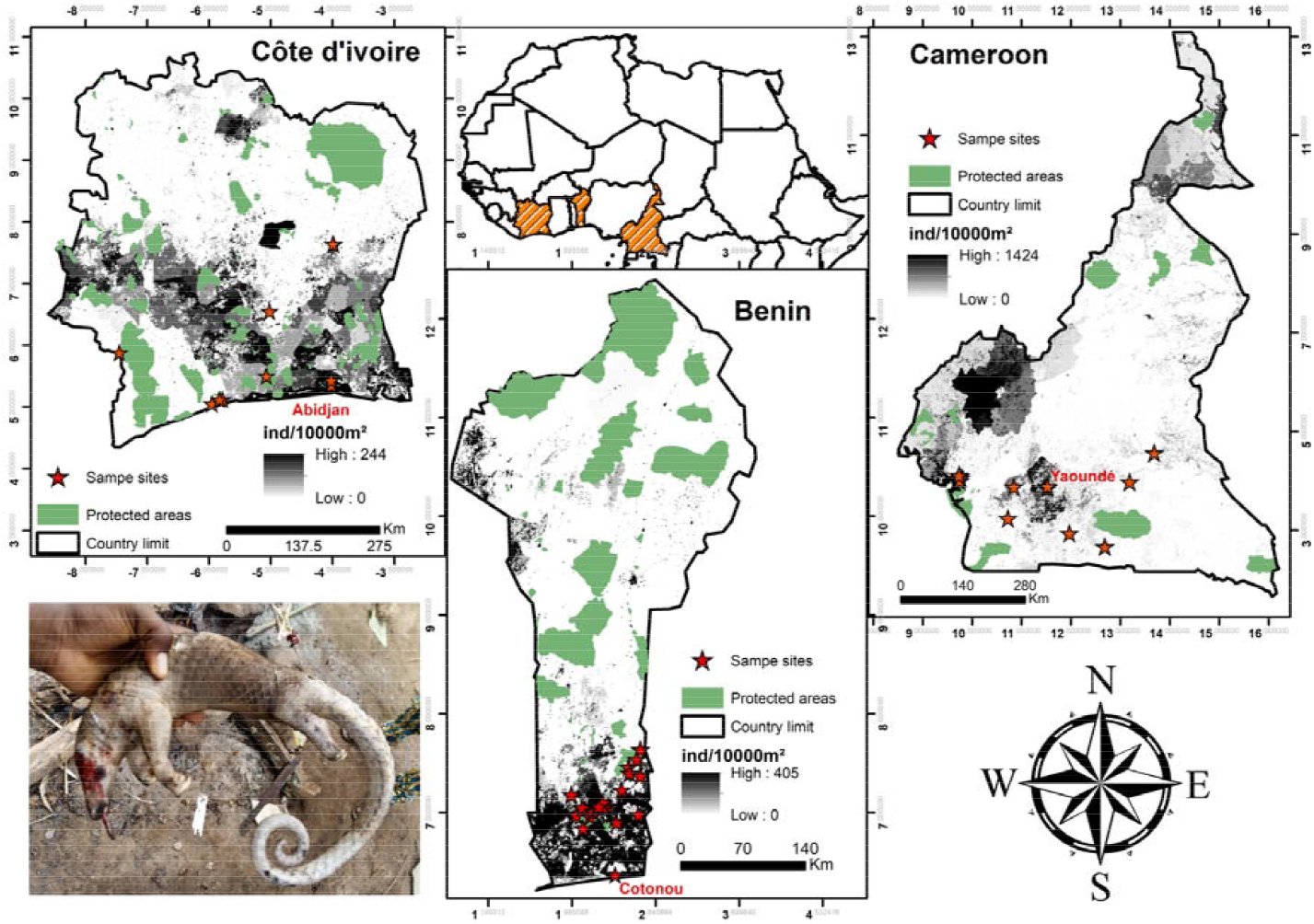
Distribution of the 48 bushmeat trade sites surveyed across West Africa as part of this study. Bottom left: a white-bellied pangolin (*Phataginus tricuspis*) traded in southern Benin (photo credits: Stanislas Zanvo).

### Questionnaire

We designed a semi-structured questionnaire (Appendix Document 1) where we first collected information on the sociodemographic status of vendors (age, gender, ethnic group, education, seniority), and then asked questions about (i) the impact of the COVID-19 pandemic on bushmeat trade activities in general, (ii) its impact on the pangolin trade in particular, (iii) the effect of the sanitary measures taken by governments, and (iv) how sanitary risks related to the bushmeat trade activities and pangolins were perceived. Interviews were conducted in French or in the local language, after recruiting translators within the bushmeat trade communities from pre-survey meetings. We explained the purposes of our study and ask for a verbal or signed consent before starting the interviews with the vendors. Each respondent was interviewed for an average of 35 minutes. Interview sheets were sometimes given to certain actors whose level of education was considered sufficient on the basis of their ability to read and understand the questions after a discussion of five to ten minutes when these latter could not grant us more time at the time of the survey. Overall, questionnaires with evasive or incomplete answers were systematically excluded.

### Descriptive and statistical analyses

In total, 32 answers to our questionnaire were analyzed. Sociodemographic variables were quantitative and qualitative. The other variables extracted from the questionnaire were derived from 20 close-ended and seven open-ended questions. For descriptive analysis of the variables, graphic quantifications were conducted in Excel 2016 using dynamic tables. XLSTAT 2021.4.1 (Addinsoft, 2022) was used for a series of statistical tests. The normality of the distribution of men’s and women’s age pyramids per country was assessed through a Shapiro-Wilk test (alpha = 0.05), and was rejected in all cases except female vendors from Benin (data not shown). Differences in distribution were compared using a non-parametric Kolmogorov-Smirnov test (alpha = 0.05). Linear regression analysis between age and seniority was conducted to assess the correlation between the two variables. We used the Χ² test –or Fisher’s exact test when answers were below 5 in number– (alpha = 0.01 in both cases) in order to assess whether answers to close-ended questions differed among the three countries. Differences in periods of cessation of trade activities reported among countries were assessed through a Wilks’ G² independency test (alpha = 0.01). Word clouds were used to summarize answers to open-ended questions.

## Results

### Socio-demographic variables

Women were dominant (63.6%) or highly dominant among the vendors (98.8%) in Cameroon and Côte d’Ivoire, respectively. In Benin, vendors were mostly men (71.7%) (Appendix Figure 1). Distributions along the age pyramids of men and women were not significantly different in Benin and Cameroon (P>0.05; D=0.500 and 0.462; Côte d’Ivoire could not be tested because of only one male vendor). In Côte d’Ivoire, 14 ethnic groups or nationalities were involved as bushmeat vendors, where *Baoulé* dominated (45%). In Benin, vendors were composed of seven ethnic groups and mostly represented by *Fon* (46%). A total of 21 ethnic groups were involved as vendors in Cameroon, with *Beti* (34%) and *Bassa’a* (18%) representing more than half of the interviewees (Appendix Figure 2). Seniority in the trade ranged from a few months to 45 years, without any apparent differences among the countries (Appendix Figure 3). Combining data from the three countries, age varied from 15 to 87 yr and was significantly correlated with seniority (R²=0.307; P<0.0001) (Fig. 2). In general, vendors from Côte d’Ivoire and Benin had no scholar education or stopped during primary school, while a college level education was preponderant in Cameroonian vendors (Appendix Figure 4).

**Figure 2.**
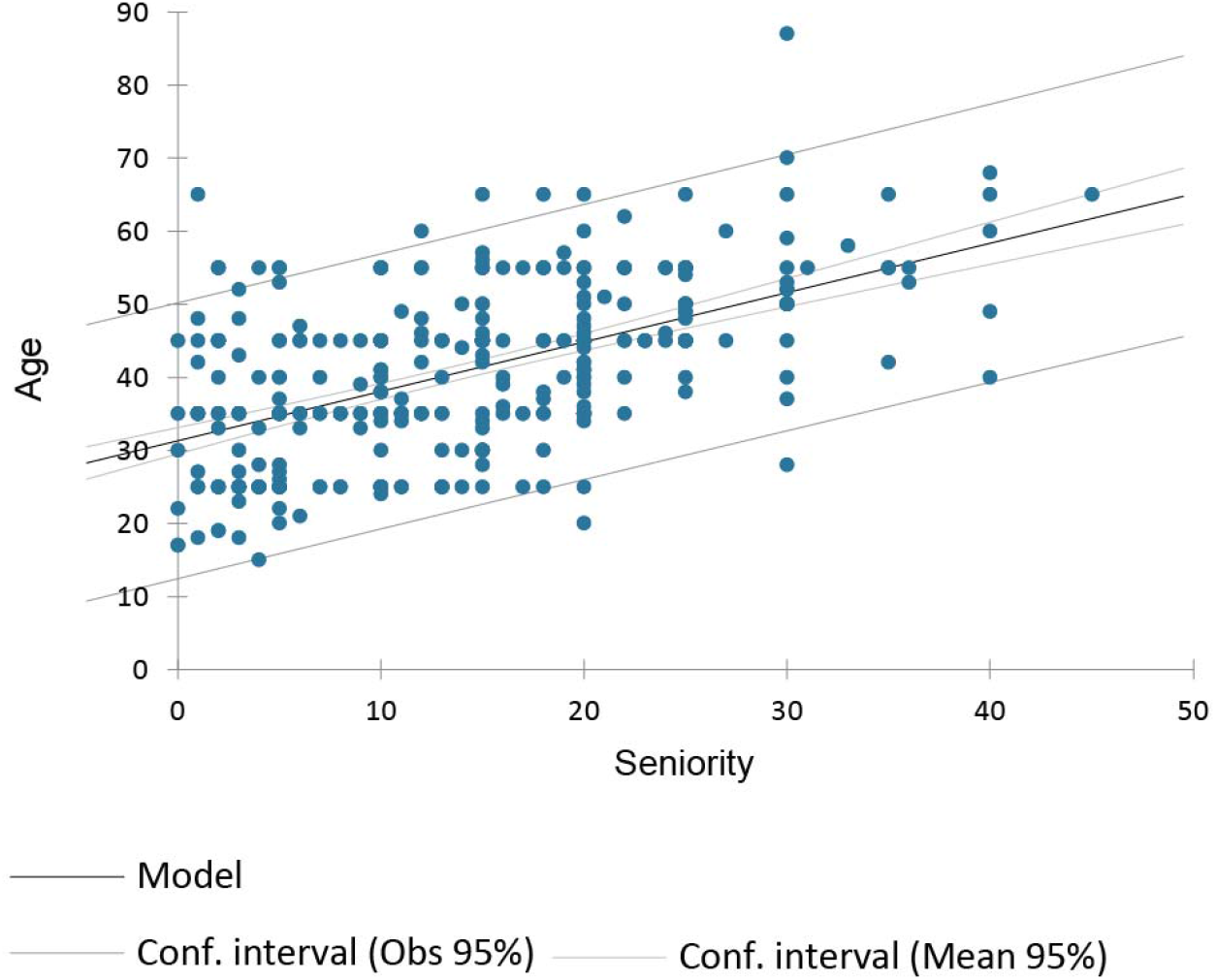
Linear regression between age and seniority (in years) of bushmeat vendors across three western African countries (Côte d’Ivoire, Benin and Cameroon). N observations = 361.

### Impact of the COVID-19 pandemic on bushmeat trade activities

Most vendors from the three countries (75.0-91.8%) said that there had been a negative impact of the COVID-19 pandemic on their activities (P=0.000; Χ²=16.583 among countries). This impact was still prevailing at the time of the study in the three countries for 68.7-78.0% of the interviewees (P=0.239; X²=2.862) and involved a global decrease in the number of clients for 73.7-89.0% of the vendors (P=0.022; X²=7.650) (Appendix Figure 5). However, vendors’ answers were significantly different among countries as to the temporary stop of their activities related to COVID-19 governmental measures (P<0.0001; X²=74.088). Most of the vendors in Côte d’Ivoire (83.0%) had to temporarily stop their activities, whereas it was not the case in Benin (44.0% of the cases) and – particularly– in Cameroon (only 28.6% of the vendors) (Appendix Figure 6). The temporary cessation of trade activities also varied in time among countries (P<0.0001; Wilks’ G²=83.565), with distributions skewed towards durations less than two months in Benin and Cameroon (67 and 80.8%, respectively), whereas Côte d’Ivoire had a larger spectrum where three months dominated (Fig. 3). Vendors from Benin mostly reported (84.0%) an impact of the pandemic on the bushmeat supply chain, whereas the picture was more mitigated in Côte d’Ivoire and Cameroon (55.8 and 53.9% of the interviewees, respectively). In Côte d’Ivoire, the selling prices of bushmeat were reported to decrease in 86.3% of the cases, while price stability was mostly mentioned in Benin (88.0%) and Cameroon (68.7%) (Appendix Figure 7). Differences in the impact on the supply chain and selling prices among countries were both significant (P<0.0001; Χ²=27.311 and 123.239). The great majority of vendors across the three countries (91.0-96.8%) reported that they will continue their activities despite the impact of the COVID-19 pandemic (P= 0.094, Fisher’s exact test) (Appendix Figure 8), as they “need money” (83.6%) (Appendix Figure 9). The bushmeat trade was declared as the only source of revenue for 68.3% of the vendors, and a direct source of income for the household (“family”; 25.5%) (Appendix Figure 10).

**Figure 3.**
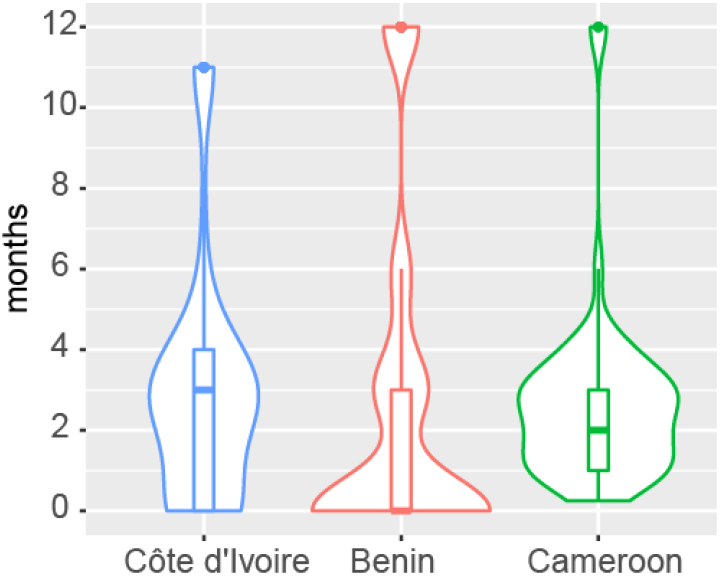
Violin plot representation of the time during which bushmeat vendors had to stop their activities in Côte d’Ivoire, Benin and Cameroon.

### Impact of the COVID-19 pandemic on the pangolin trade

Most of the vendors recognized the negative impact of the COVID-19 pandemic on the pangolin trade in Benin (68.0%) and Cameroon (81.3%), whereas the picture was more balanced in Côte d’Ivoire (51.6%) (P<0.0001; Χ²=26.779). Benin and Cameroon mostly reported a decrease in the number of clients for pangolins (68.0 and 67.6%, respectively), whereas Côte d’Ivoire did not (13.7%) (P<0.0001; X²=83.235) (Appendix Figure 11). Most vendors from Benin (66.0%) notified difficulties in the supply chain for pangolins, while such difficulties were reported by approximately half of the vendors from Côte d’Ivoire (44.2%) and Cameroon (51.7%) (P=0.009; X²=9.469). Stockpiling pangolins as a solution against difficulties with the supply chain was rarely considered in Côte d’Ivoire (13.7%) and Benin (7.0%), whereas 42.9% of the vendors used stockpiling in Cameroon (P<0.0001; X²=53.140) (Fig. 4). The selling prices of pangolins were mostly diminished in Benin (70.0%) and Cameroon (58.8%), while they were almost not impacted in Côte d’Ivoire (7.4%) (P<0.0001; X²=90.951) (Appendix Figure 12).

**Figure 4.**
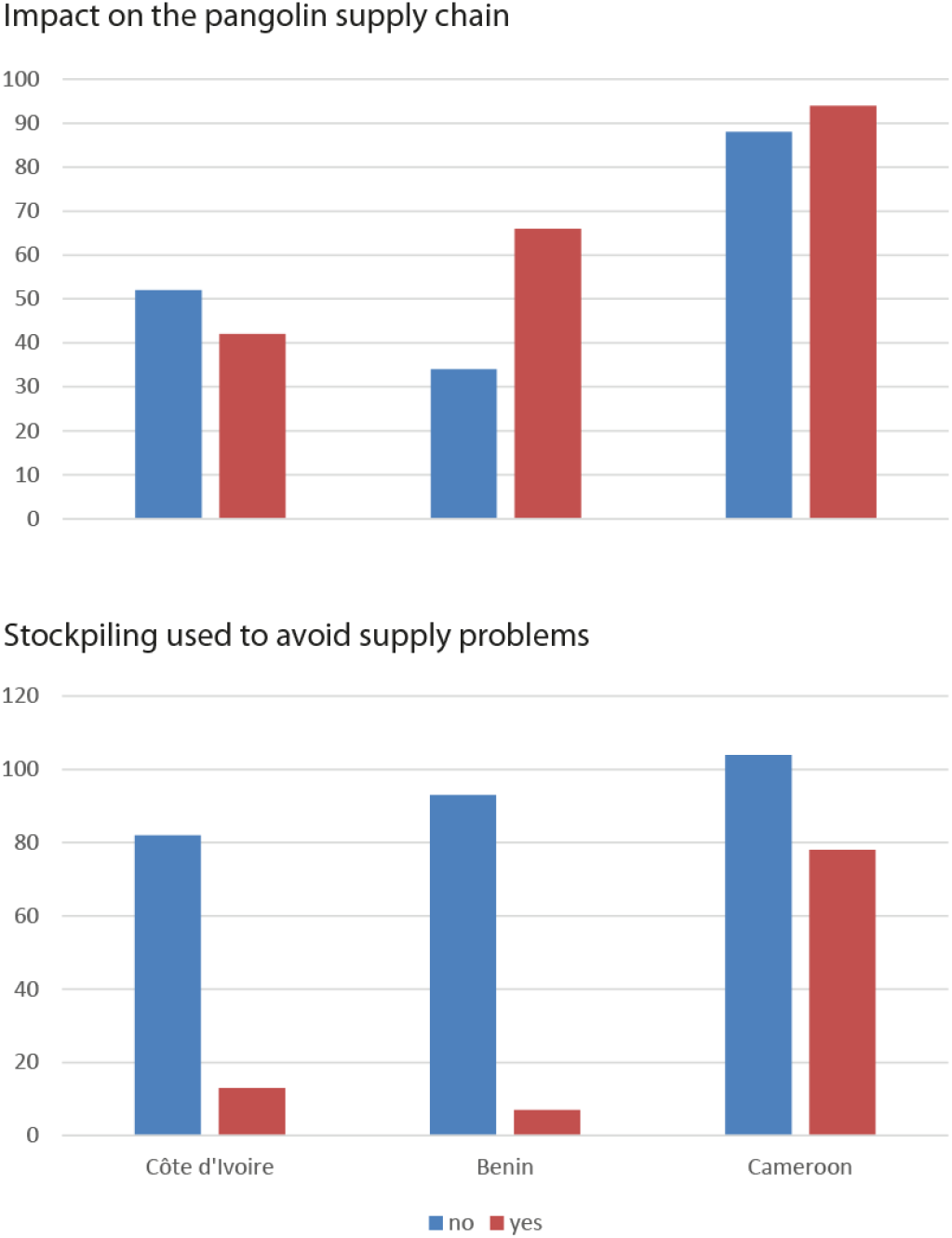
Impact of the COVID-19 pandemic on the availability of pangolins in bushmeat markets as perceived among vendors from Côte d’Ivoire, Benin and Cameroon. Top: negative impact on the pangolin trade supply chain. Bottom: stockpiling counter strategy.

### Perceptions of the sanitary measures taken by governments

Most vendors from Côte d’Ivoire (63.1%) and Benin (84%) reported governmental measures for controlling the bushmeat markets, whereas almost only half of the vendors from Cameroon (52.8%) did (P<0.0001; X²=27.263) (Appendix Figure 13). The reported governmental actions were mostly protective measures against the COVID-19 pandemic in Benin (84.4%) and Cameroon (56.0%), while the ban on the bushmeat trade was mostly cited by vendors from Côte d’Ivoire (64.7%; cumulating 93.1% with protective measures) (Fig. 5). Perceptions on governmental measures were generally good in Benin (62.5%; no opinion: 26.0%) and Cameroon (48.9%; no opinion: 23.6%), but were mostly seen as negative in Côte d’Ivoire (56.8%; no opinion: 12.6%) (P<0.0001; X²=48.322) (Appendix Figure 14). Globally, the impact of the governmental measures on trade activities was seen as deleterious among countries (“bad”: 32.0%; “less clients”: 30.6%), with only 4.8% of the vendors considering these measures as “good for safety” (Appendix Figure 15).

**Figure 5.**
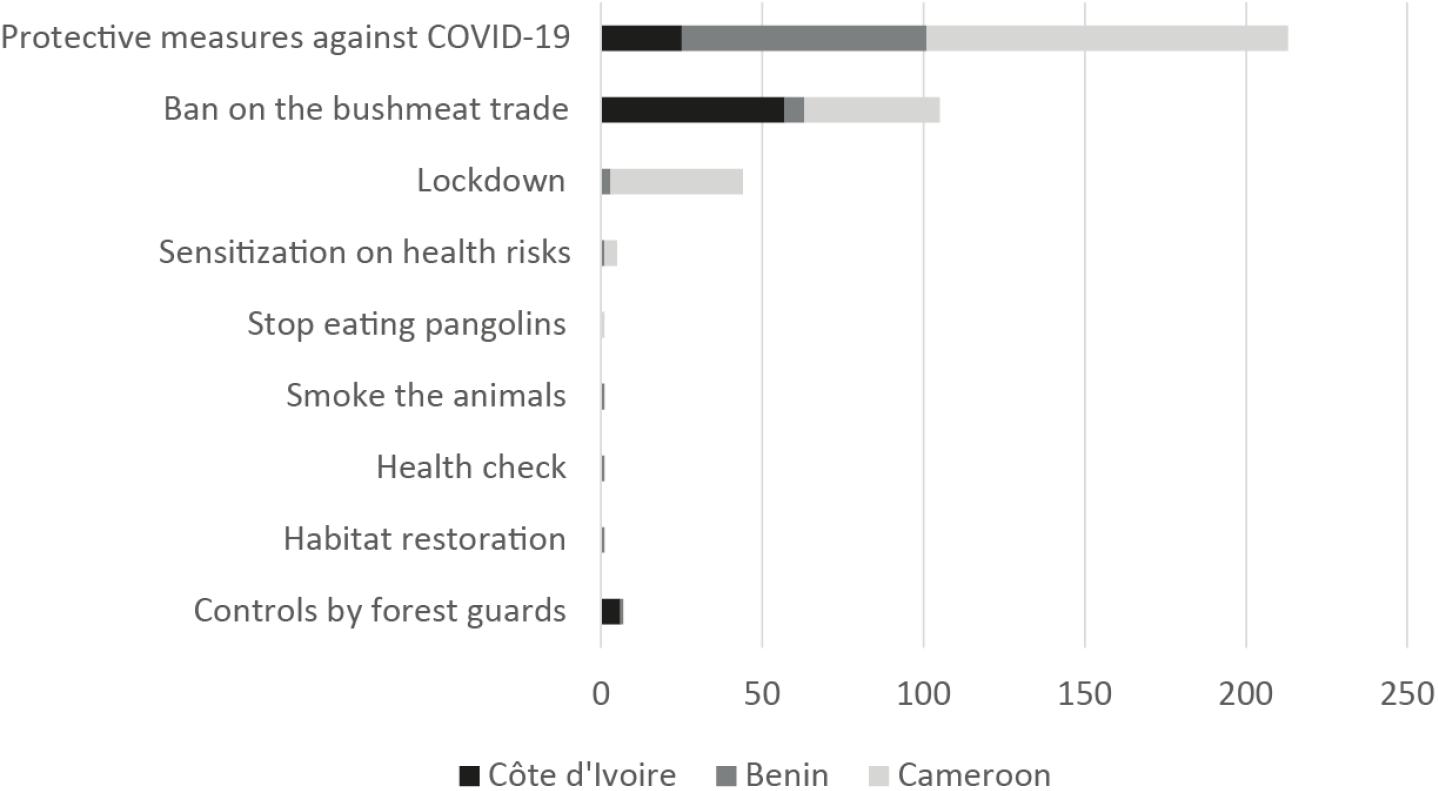
Governmental measures against the COVID-19 pandemic as described by bushmeat vendors from Côte d’Ivoire, Benin and Cameroon.

### Perceptions of sanitary risks related to the bushmeat trade activities and pangolins

The majority of the vendors in Benin (61.0%) and Cameroon (62.6%) mentioned their awareness of the sanitary risks related to the bushmeat trade and of diseases linked to their activities (73.0 and 66.5%, respectively), whereas in Côte d’Ivoire most of the vendors said that they were not aware of such risks (75.8%) and diseases (76.8%) (P<0.0001 in both cases; X²=40.886 and 64.829) (Appendix Figure 16). Ebola virus disease was mentioned as the main potential disease in most cases across the three countries (59.3-87.4%), with Lassa fever only reported in Benin and at an important proportion (30.9%) (Appendix Figure 17). In their majority, vendors generally did not report being sick in relation to their activities (from 6.08% in Cameroon, to 35.0% in Benin and 45.3% in Côte d’Ivoire) (P<0.0001; X²=60.754) (Appendix Figure 18). Cold with fever and/or cough was mostly reported from Cameroon (81.7%; but total number of answers = 11). Malaria came first in Côte d’Ivoire (78.7%) and Benin (44.1%), Benin also cumulating cold with cough (35.3%) (Appendix Figure 19).

Knowledge on the potential role played by pangolins in the COVID-19 pandemic was high in Cameroon (82.4%), but moderate in Côte d’Ivoire (51.0%) and Benin (46.3%) (P<0.0001; X²=47.455) (Appendix Figure 20). The main sources of information used to access knowledge on the role of pangolins were television (25.5%) and social networks (22.4%) (Fig. 6). Most of the vendors did not believe in the information reporting the link between pangolins and COVID-19 (76.0 to 90.1% of the interviewees per country) (Appendix Figure 21). The main reasons were because “people have always been eating pangolins and are not sick” (55.1%) and “it is fake information” (19.7%). On the other hand, “media cover” was taken as the best guarantee for the veracity of the information in vendors whom believed in the link between pangolins and COVID-19 (28%) (Fig. 6).

**Figure 6.**
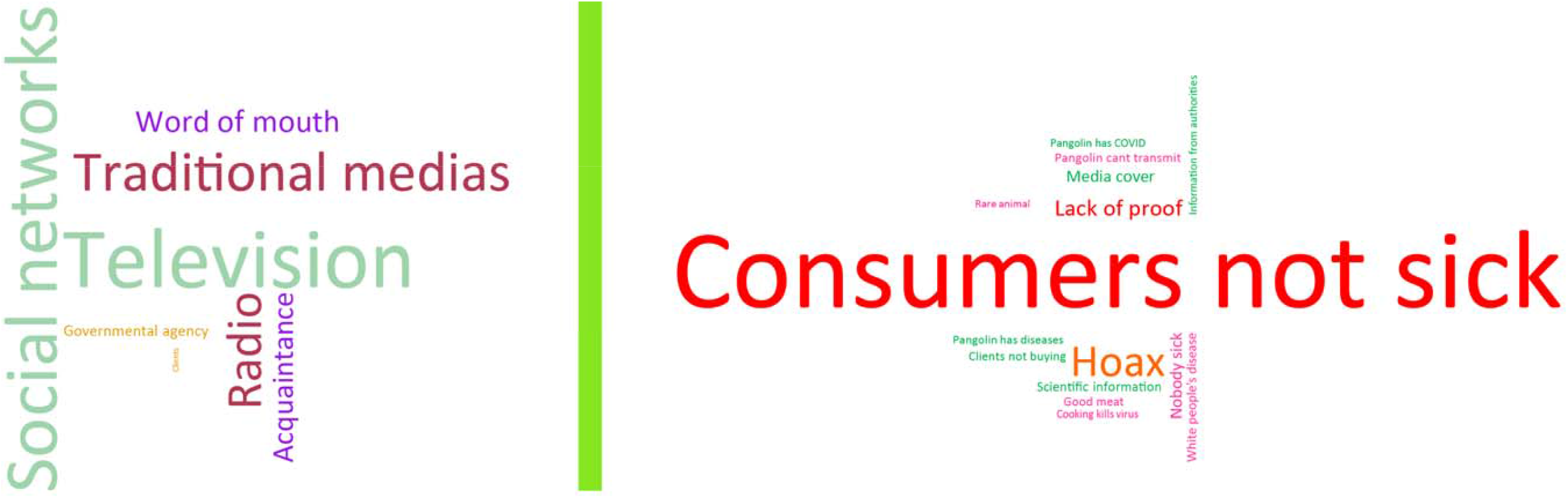
Word cloud representation of the sources of information used by vendors (left) and the reasons why they believe or not in the role of pangolins in the COVID-19 pandemic (right). Right: words have been colored in red-to-pink tones for answers justifying why vendors did not believe in the role of pangolins in the COVID-19 pandemic, and in green tones for answers justifying why vendors believed in the role played by pangolins. Word size is correlated to the number of times the answer was given. Example: Television – 74 times (left); Consumers not sick – 162 times (right).

## Discussion

Our unprecedented comparative survey encompassing three West African countries allowed revealing some major sociodemographic patterns among bushmeat vendors. Because bushmeat surveys have mostly been implemented at the local scale (Groom et al., in prep.), the global picture of ethnic contributions to the national bushmeat trade has never been properly assessed (but see Randolph et al. 2022). We found that one major ethnic group was dominating the trade in each country, involving *Baoulé* in Côte d’Ivoire, *Fon* in Benin and *Beti*-*Bassa’a* in Cameroon (within a wide diversity of ethnic groups involved in each country). Zanvo et al. (2021b) recently suggested that *Fon* were the dominant and most anciently involved ethnic group in the wildlife trade in Benin. It is possible that the dominance of single ethnic groups across national bushmeat trade networks is due to ancestral stranglehold on the market, subsequently seated by transmitted systems of rules through cultural group selection (see Richerson et al. 2016), still operational today. However, the ethnic patterns that we described should be taken with caution, as our survey efforts were geographically biased towards the southern region of each country (where the bushmeat trade thrives).

We observed contrasting patterns in gender distribution among bushmeat vendors, from women being highly dominant in Côte d’Ivoire (and dominant in Cameroon, in line with Randolph et al. 2022) to men being preponderant in Benin. Mixed or female-biased gender contributions have largely been reported across West and central Africa (e.g., Kamins et al. 2015; Lucas et al. 2022), whereas male-biased distribution may be a particularity of Benin, where the wildlife trade is dominated by traditional medicine markets rooted into the *Vodun* practices (Zanvo et al. 2021a). Gendered structures could be important to consider in the bushmeat trade dynamics as, together with ethnic group selection, they may lead to differentiated responses to a same constraint. Whether gender influenced perceptions on the measures against the COVID-19 pandemic was not directly tested in our study but could be further investigated using our database and future surveys.

One striking pattern was the significant, positive correlation between age and seniority among the bushmeat vendors. To our knowledge, this is the first time that a study demonstrates on a large scale (West Africa) that being a bushmeat trade vendor means engaging into a stable career. Such a pattern may be at least partly explained by the overall low level of education observed among vendors, low education –coupled with family- and/or ethnicity– fostering the will to build long-term careers in the bushmeat trade (Randolph et al. 2022).

There was a general consensus among bushmeat vendors from West Africa that the COVID-19 pandemic and related governmental measures had a negative impact on their activities, as translated into a decrease in number of clients. Although information on the economic impact of the COVID-19 pandemic on the bushmeat trade remains extremely scarce, governmental measures in Nigeria seem to have significantly decreased the volumes of bushmeat sold on the markets (Funk et al. 2022). Such impact was still perceived as visible by most of the interviewed West African vendors after the end of governmental measures against the pandemic. This is supported by the partial recovery (c. 56 to 91%) from initial numbers of bushmeat vendors in Côte d’Ivoire as observed eight months after the end of the lockdown (Gossé et al. in prep.). During the Ebola epidemic in 2014, a prolonged impact of the governmental ban was also observed as post-epidemic recovery in bushmeat volumes was not achieved for some taxa (Funk et al. 2021). However, bushmeat trade dynamics may not be fully captured by either vendors’ interviews or bushmeat and vendors’ counts alone. Indeed, online trade –as operating in West Africa (Moloney et al., in prep.)– can constitute a loophole to the governmental constraints imposed on the wildlife trade (Morcatty et al. 2021).

Beyond that global consensus, we observed significantly differentiated appreciations of the impact of the COVID-19 pandemic among West African vendors. Côte d’Ivoire was hit by more severely perceived governmental measures, specifically translated into a stricter and longer ban (median around three months) compared to Cameroon (< 2 months) and Benin (around zero). As a corollary, the selling prices of the bushmeat were reported to decrease by most Ivorian vendors whereas they were generally estimated to remain stable in Benin and Cameroon, where governmental actions were mostly reported as protective measures against the COVID-19. Côte d’Ivoire occupies a special place in the West African panorama, as hunting for bushmeat has been officially banned from the country since 1974 (Gonedelé-Bi et al. 2022). Although the bushmeat trade remains widespread and openly accessible (Gossé et al. 2022), we believe that the cumulated effect of such national ban and the sensitization and repression measures operated during the 2014 Ebola pandemic (World Health Organization 2015; Dindé et al. 2017), may have promoted the greater implementation by Ivorian authorities of COVID-19 mitigation measures.

This situation was seriously contrasting with that observed in Cameroon, where only c. 1/4 of the vendors reported a temporary stop of their activities, and were the lowest in numbers (c. 53%) to report that governmental actions took place. This is in line with the fact that actions from the successive Cameroonian governments to mitigate the bushmeat trade have so far been hampered by state corruption (Peh and Drori 2010) or resulted in shifting places where trade operates (Randolph 2016), ultimately jeopardizing measures to mitigate the COVID-19 pandemic. In Benin, where c. 60% of the vendors did not experience a stop in their activities, the contrast with Côte d’Ivoire is even stronger. The Beninese wildlife trade is organized between bushmeat and traditional medicine markets (Djagoun et al. 2013). Traditional medicine markets being predominant and deeply anchored into the religious culture of the country, it is possible that for political reasons, governmental controls were less strictly applied at the time of the lockdown. Another potential reason for this relaxed impact of national sanitary measures observed in Benin and Cameroon is that both countries have so far been spared by Ebola outbreaks (N’koué Sambiéni et al. 2015; Wirsiy et al. 2021).

Significantly differentiated appreciations also prevailed as to the impact of the pandemic on the bushmeat supply chain, which was strongly reported as negatively affected in Benin –including for pangolins– compared to Côte d’Ivoire and Cameroon, where the picture was more mitigated. Benin is a small country with a bushmeat supply chain endemic to the Dahomey Gap (see Zanvo et al. 2022 for an example on the pangolin trade), a narrow savannah-forest mosaic zone where sourced forests are isolated and highly fragmented (Salzmann and Hoelzmann 2005). Given this socio-geographical context, it is expected that lockdown measures restricting movements, even light, could have significantly impacted the supply chain in the country.

The fact that vendors from Côte d’Ivoire, although acknowledging a strong ban on their bushmeat trade activities, did not massively report a negative impact of the COVID-19 pandemic on the supply chain may sound counterintuitive. All the more since Ivorian vendors also concluded to a mitigated-to-shallow impact on the pangolin trade, in terms of number of clients, supply chain and selling prices. A possible explanation could be that the Ivorian market is fed by a wide spectrum of bushmeat sites, especially in the case of urban centers (Gossé 2023), thus providing a wealth of alternative options to skirt mitigation measures once the ban ended. Stockpiling pangolins during the ban period could have also been a response to counter difficulties with the supply chain (see Emogor et al. 2021), but this strategy was only reported in fair numbers (c. 43% of the vendors) in Cameroon. There and in Benin, most vendors acknowledged the negative impact of the COVID-19 pandemic on the pangolin trade, echoing the lower numbers of pangolins sold in Liberia and Cameroon during the COVID-19 pandemic (Deemie et al. 2021; Harvey-Carroll et al. 2022).

Eventually, our results showed that the bushmeat trade constituted the main source of wealth for most of the vendors across West Africa, and that for this reason they would continue their activities despite the existence of mitigating measures such as those related to the COVID-19 pandemic. Although the economic contribution of the bushmeat trade-related incomes to rural households has been largely –and with contradictory conclusions– investigated (Nielsen et al. 2017), the question of the profitability of the trade to end-point vendors remains poorly explored. Despite low expected realized margins (Cowlishaw et al. 2005), we show that the carrier of bushmeat vendors is generally perceived as profitable enough to constitute a long-term, main activity, notably in urban centers. As such, we recognize bushmeat vendors as a socio-professional category that will be decisive to include in strategies intended to mitigate or control bushmeat hunting networks.

We reported significant differences in the perceptions of sanitary risks and governmental measures among West African bushmeat vendors in relation to their activities. A majority of vendors from Benin and Cameroon mentioned their awareness of sanitary risks and related diseases, and considered the governmental measures (mostly against COVID-19 transmission) as positive. Ebola virus disease, together with Lassa fever in Benin, were cited as the main diseases linked to the bushmeat trade. Those two deadly zoonoses have been well studied in West Africa (e.g., De Nys et al. 2018; Yadouleton et al. 2020) and awareness campaigns may have been successful in some countries (Nelson and Namtira 2017; Odionye et al. 2019). However, awareness of the risks and diseases is not equivalent to recognition by the actors of the genuine level of sanitary risks related to their activities (Saylors et al. 2021). As a matter of fact, vendors from Benin and Cameroon poorly reported being sick during their periods of activity. The symptoms given could not be related to any specific zoonoses, as the major responses (cold with fever or cough) could apply to anything from a simple cold to malaria, including COVID-19.

On the other hand, low awareness among Ivorian vendors of the diseases transmitted by the bushmeat was surprising (as was the greatest number of vendors mentioning being sick in relation to their activities), given the important awareness campaigns that occurred during the 2014 Ebola epidemic (World Health Organization 2015). The predominant negative perceptions of governmental measures in Côte d’Ivoire is likely explained by a combination of factors, including low awareness of sanitary risks related to the bushmeat trade and the repressive context in place since the trade ban that occurred during the Ebola outbreak (Dindé et al. 2017).

Knowledge on the potential role played by pangolins in the COVID-19 pandemic was significantly variable in West Africa, being much higher in Cameroon than in Côte d’Ivoire and Benin. The main sources of information were identified as television and social networks, two media that have diffused a range of contrasting and sometimes poorly informative information on the nature of the COVID-19 pandemic, notably in Cameroon (Djofack and Bien A Ngon 2020). Nevertheless, such media were trusted as a guarantee of information accuracy by 28% of the vendors who believed in the role played by pangolins. Media can be an alternative, powerful tool to compensate for state weaknesses and citizens’ lack of trust in the context of a pandemic (Odionye et al. 2019). However, several surveys on the recent epidemic of Lassa fever in West Africa have emphasized the need for increased intensity and improved accuracy of awareness campaigns in the media in order to reach genuine awareness and impact behaviors (Wogu 2018; Wogu et al. 2020). Such recommendation also applies to the COVID-19 situation in Africa (Okereke et al. 2020).

A massive proportion of West African vendors did not believe that pangolins could be involved in the pandemic. Far beyond the *fake origin* hypothesis of the COVID-19 pandemic emitted by part of the vendors, the main reason was that “people have always been eating pangolins and have never been sick”. Traditional knowledge is not necessarily a safeguard against the damageable use of wildlife, both for humans and the ecosystems (Alves and Rosa 2005). Here, the immutable vision for the safe consumption of pangolins as expressed by most of the West African vendors, is in serious contradiction with the evolutionary nature of life, and in this particular case, viruses. Indeed, recombination and positive selection along viral genomes can quickly turn a harmless viral strain into an emerging infectious disease (Forni et al. 2017; Visher et al. 2021), as was likely the case for the SARS-CoV-2 at the origin of the COVID-19 pandemic (Singh and Yi 2021). Beyond standard sensitization on the paths of zoonotic transmission, we suggest that better compliance with sanitary measures should also rely on educating bushmeat actors on the nature of microbial evolution.

## Conclusion

Our study showed that bushmeat trade vendors across West Africa constitute a socio-economic category driven by ethnicity and (generally) gender bias. This suggests that compliance and resilience to national sanitary measures will at least partly depend on the socio-demographic aspects proper to each national market network. We also demonstrated that the bushmeat trade constituted the main source of wealth for most of the vendors in Côte d’Ivoire, Benin and Cameroon, being sufficiently profitable to engage into long-term careers. In view of these results, we recommend that future mitigation measures of the bushmeat trade, whether conservation- or health-oriented, take into account the national specifics of the motives and sociality of the vendors’ network driving the trade.

There was a general consensus among bushmeat vendors that the COVID-19 pandemic and related governmental measures had a negative impact on their activities and the number of clients, a cost still perceived as visible at the time of the survey. However, we observed large discrepancies among the reactions of the national markets relative to the constraints of the pandemic. Côte d’Ivoire was hardly hit by the bushmeat ban and perceived governmental measures as rather negative, whereas Cameroon generally did not report a temporary stop of the bushmeat activities (as was also the case in Benin) and engaged into the stockpiling of pangolins, and Benin suffered from a weakened supply chain. Because such differences are likely rooted in the geography and long-term political agenda of each country, predicting the impact of bushmeat trade bans and related mitigation measures on the global dynamics of bushmeat markets might be an unrealistic task if national specificities are not studied and taken into account.

Bushmeat vendors mostly received health-related information from television channels and social networks, and relied on the fixist assumption that if a species has never been a disease-vector, then it will never become one. In the case of awareness campaigns targeting the link between emerging infectious diseases and the consumption and trade of bushmeat, media will have to be better monitored in order to increase true knowledge (*vs. fake* information) on zoonotic risks and transmissions, notably by including the reasoning behind microbial evolution and host shift.

Our study revealed a complex pattern of dynamics and perceptions across bushmeat markets and vendors from different West African countries. We conclude that understanding the dynamics of the bushmeat trade will require a multidisciplinary approach where socio-economic, political, health and conservation aspects will be scrutinized together as part of long-term surveys where observation data are collected. We also recommend not to over-generalize on the global consequences of bans and governmental measures on the bushmeat trade, as each nation –the unit of our comparative study– showed specific reactions to the COVID-19 pandemic.

Eventually, we identify the socio-economic category of bushmeat vendors as a crucial point of entry of the bushmeat commodity chain. Indeed, national bans have shown that freezing this point of entry could have the power to stop the trade. Thereby, whenever efficient mitigation measures of the bushmeat trade –for sanitary or conservation purposes– need to be implemented, vendors should be a high priority target.

## Data Availability

All data produced in the present work are contained in the manuscript

## Acknowledgments

Fieldwork in Côte d’Ivoire, Benin and Cameroon, and the co-authors PG, SGB, CD, ADM, and AA, were supported by ANRS-MIE (AFRICoV – ANRS COV6). KJG and SGB were also supported by the Ministère de l’Enseignement Supérieur et de la Recherche Scientifique (MESRS) of Côte d’Ivoire, as part of the “Trace-Brousse” research project (Franco-Ivorian Cooperation within the framework of the Second Contract of Debt Reduction-Development Contracts – C2Ds) and National Research Foundation (South Africa) grant UID:133615/Nr: COV19200617532886. KJG received a PhD scholarship from ”Appui à la Modernisation et à la Réforme des Universités et Grandes Ecoles de Côte d’Ivoire” (AMRUGE-CI_N°2). PG and AA received funding from BUSHRISK (FCT IC&DT 02/SAICT/2017—n° 032130). PG also received funding from the IRD – Department ECOBIO.

## Author contributions

Philippe Gaubert: Conceptualization; Data curation; Formal analysis; Funding acquisition; Investigation; Methodology; Project administration; Resources; Software; Supervision; Validation; Visualization; Roles/Writing - original draft; Writing - review & editing. Chabi A.M.S. Djagoun, Sery Gonedelé-Bi, Alain Didier Missoup, Agostinho Antunes: Conceptualization; Funding acquisition; Methodology; Project administration; Resources; Supervision; Roles/Writing - original draft; Writing - review & editing. Maurice Tindo: Project administration; Supervision; Writing - review & editing. Koffi Jules Gossé, Cécilia Espérence Koffi, Yves Noma Noma, Edwidge Michèle N’Goran, Stanislas Zanvo, Joël Djagoun, Nazif Ales, Claude Vianney Amougou and Alain Din Dipita: Methodology; Material preparation and data collection. All authors commented on previous versions of the manuscript. All authors read and approved the final version of the manuscript.

## Figure legends

**Appendix Table 1**. Database of the questionnaire-based survey conducted among 377 vendors from 48 bushmeat trade sites in Côte d’Ivoire, Benin and Cameroon. All the variables and answers were obtained from the semi-structured questionnaire available as Appendix Document 1.

Dates, ages and ethnicity have been removed to preserve anonymity.

[Excel file]

**Appendix Figure 1.**
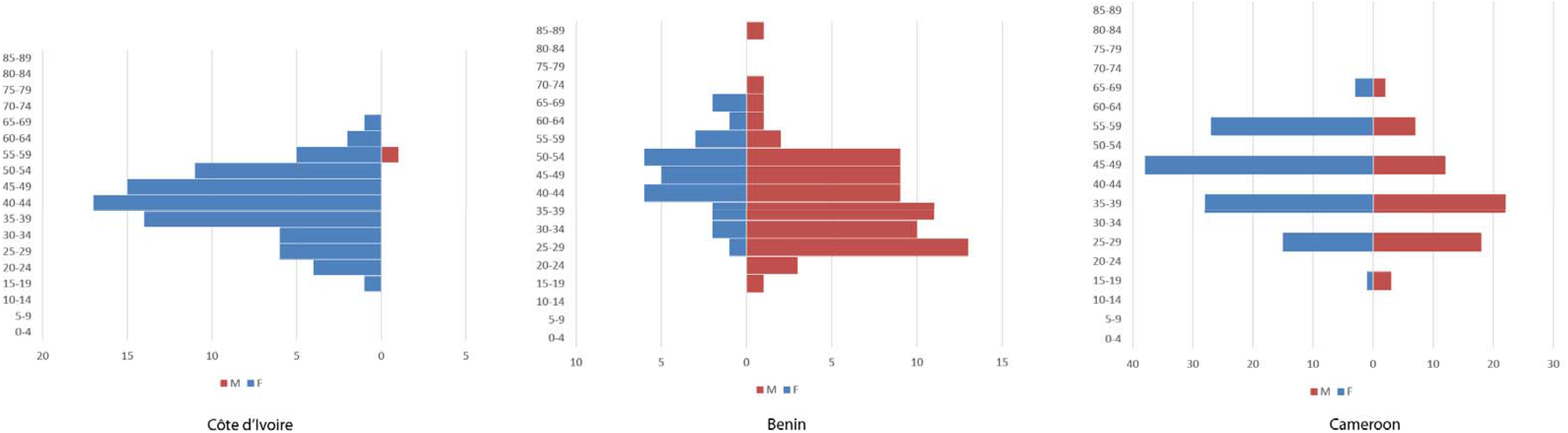
Age pyramids (in years) of the bushmeat vendors surveyed in Côte d’Ivoire, Benin and Cameroon. M = male vendor. F = female vendor.

**Appendix Figure 2.**
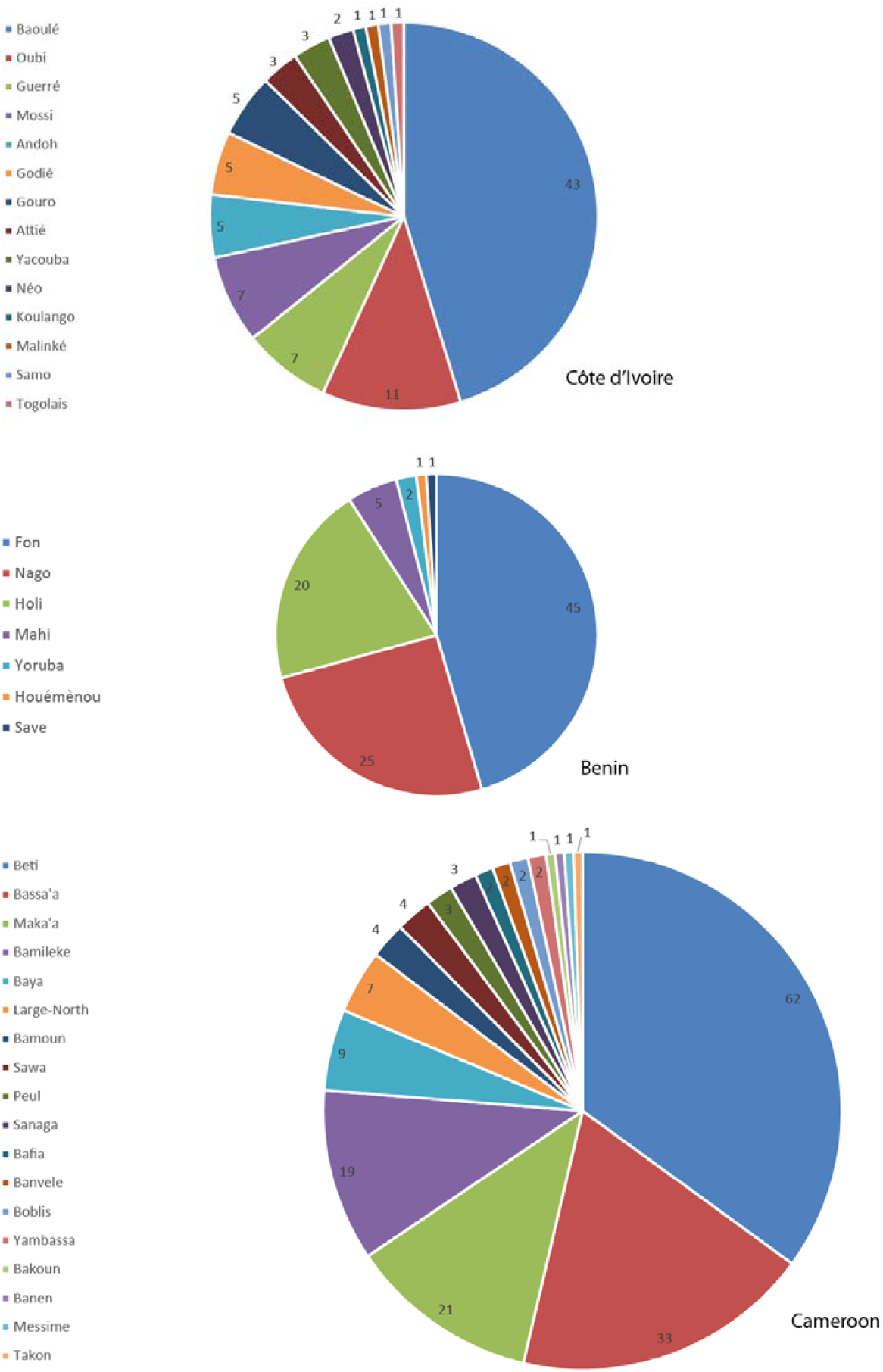
Distribution of ethnic groups among the bushmeat vendors surveyed in Côte d’Ivoire, Benin and Cameroon.

**Appendix Figure 3.**
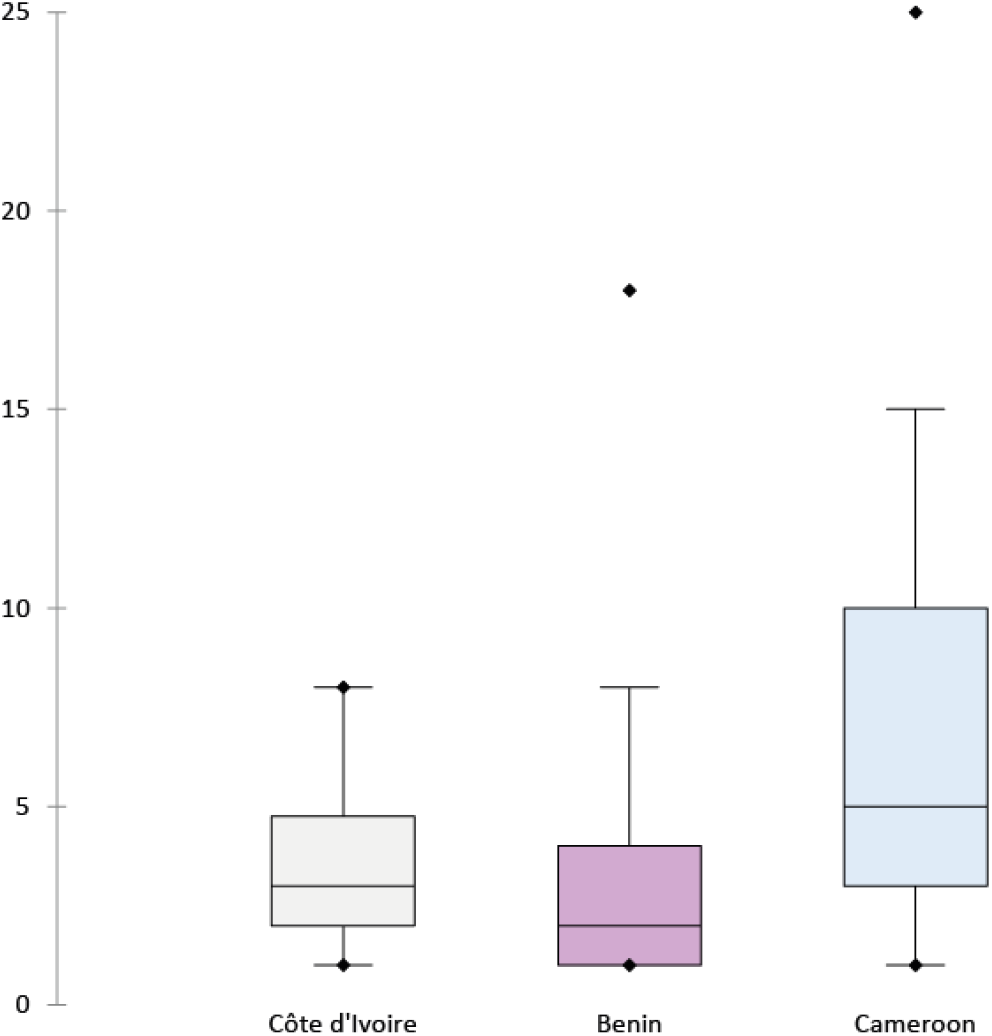
Boxplot representation of seniority in the trade of the bushmeat vendors surveyed in Côte d’Ivoire, Benin and Cameroon.

**Appendix Figure 4.**
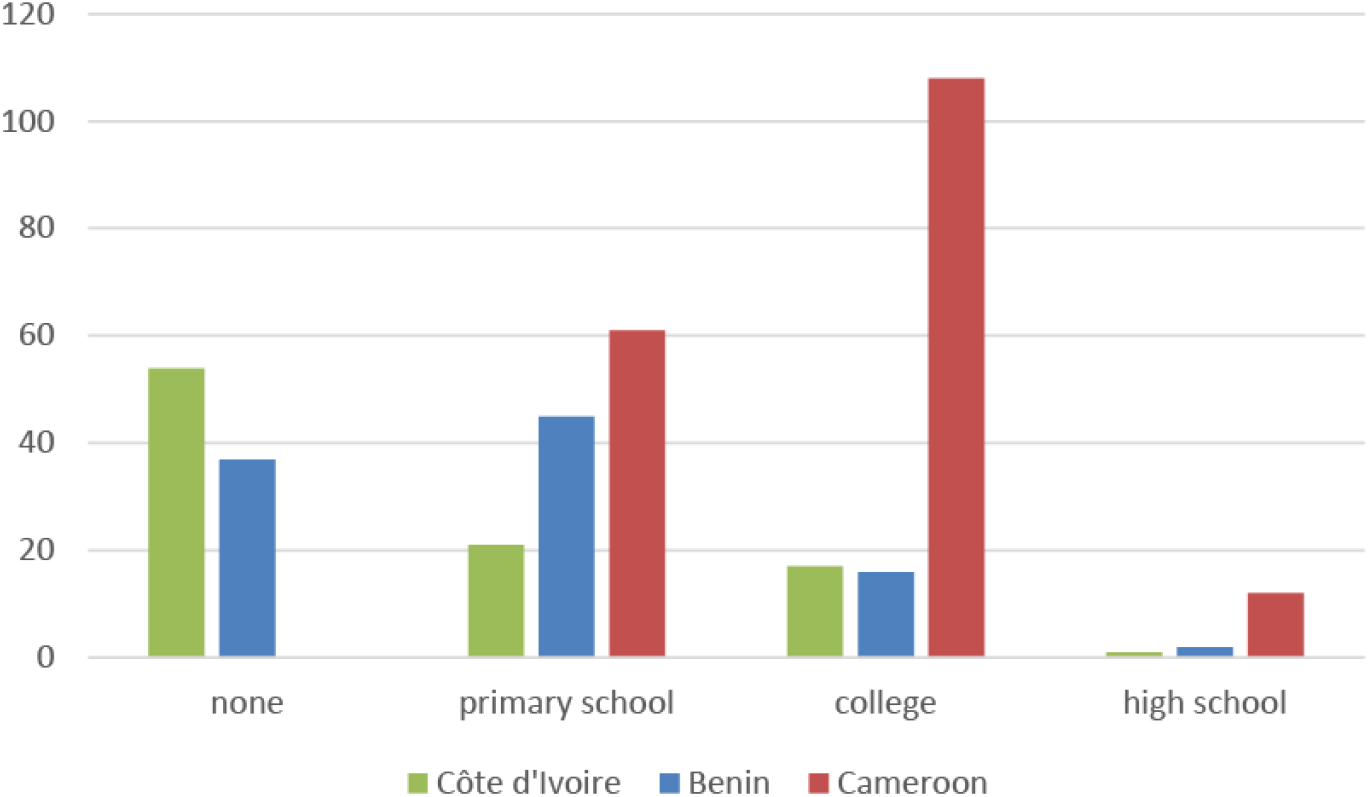
Levels of education of the bushmeat vendors surveyed in Côte d’Ivoire, Benin and Cameroon.

**Appendix Figure 5.**
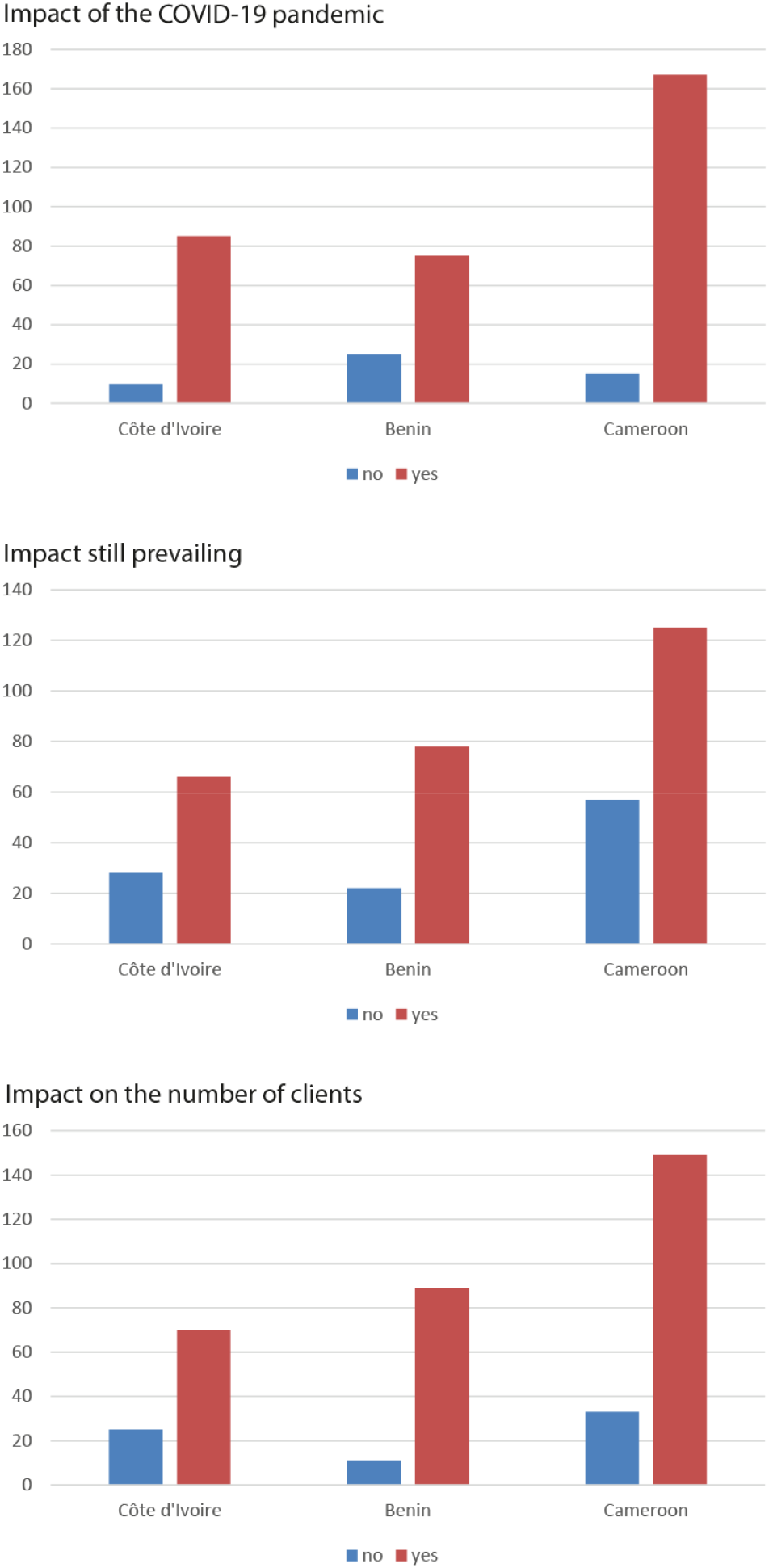
Perceived (negative) impacts of the COVID-19 pandemic on the bushmeat trade dynamics among the vendors surveyed in Côte d’Ivoire, Benin and Cameroon.

**Appendix Figure 6.**
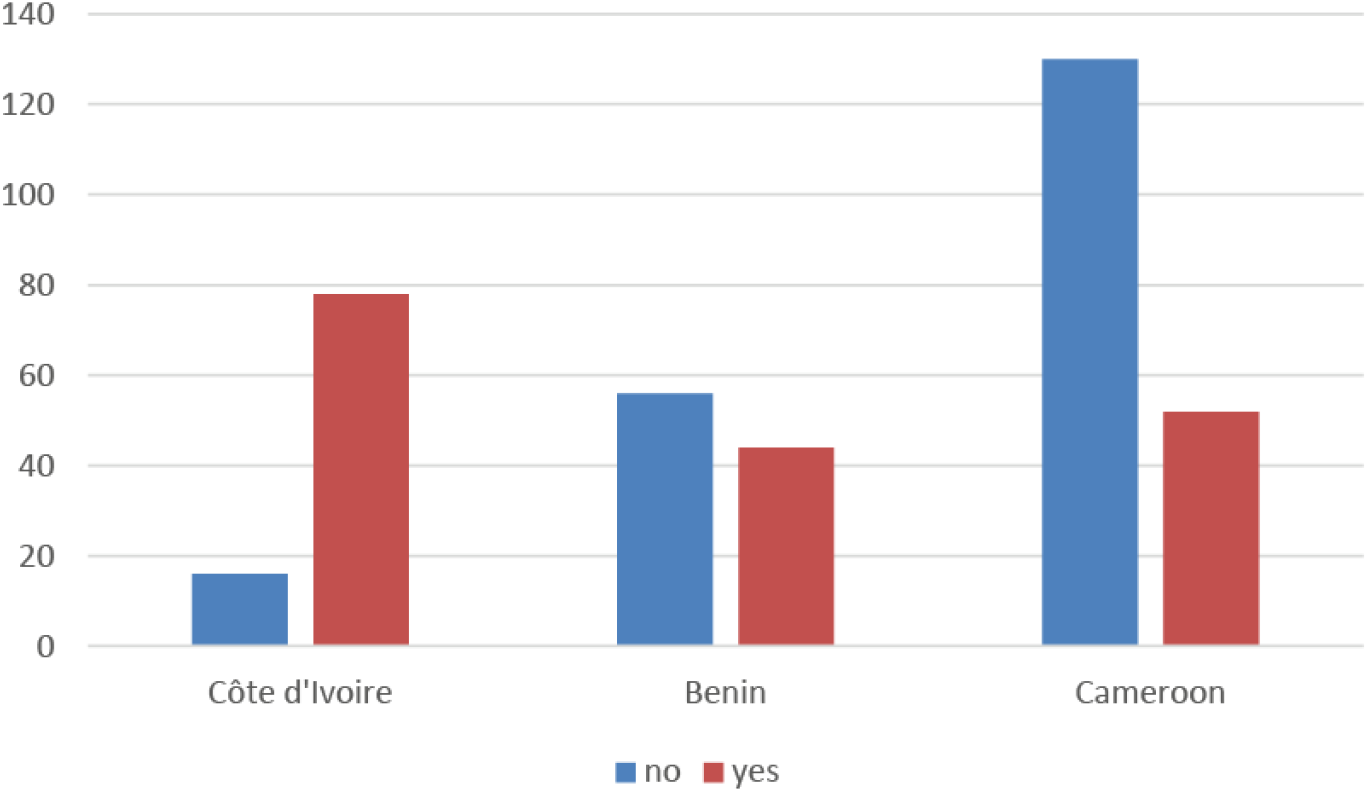
Number of vendors reporting on whether they had to stop their activities during the COVID-19 pandemic in Côte d’Ivoire, Benin and Cameroon.

**Appendix Figure 7.**
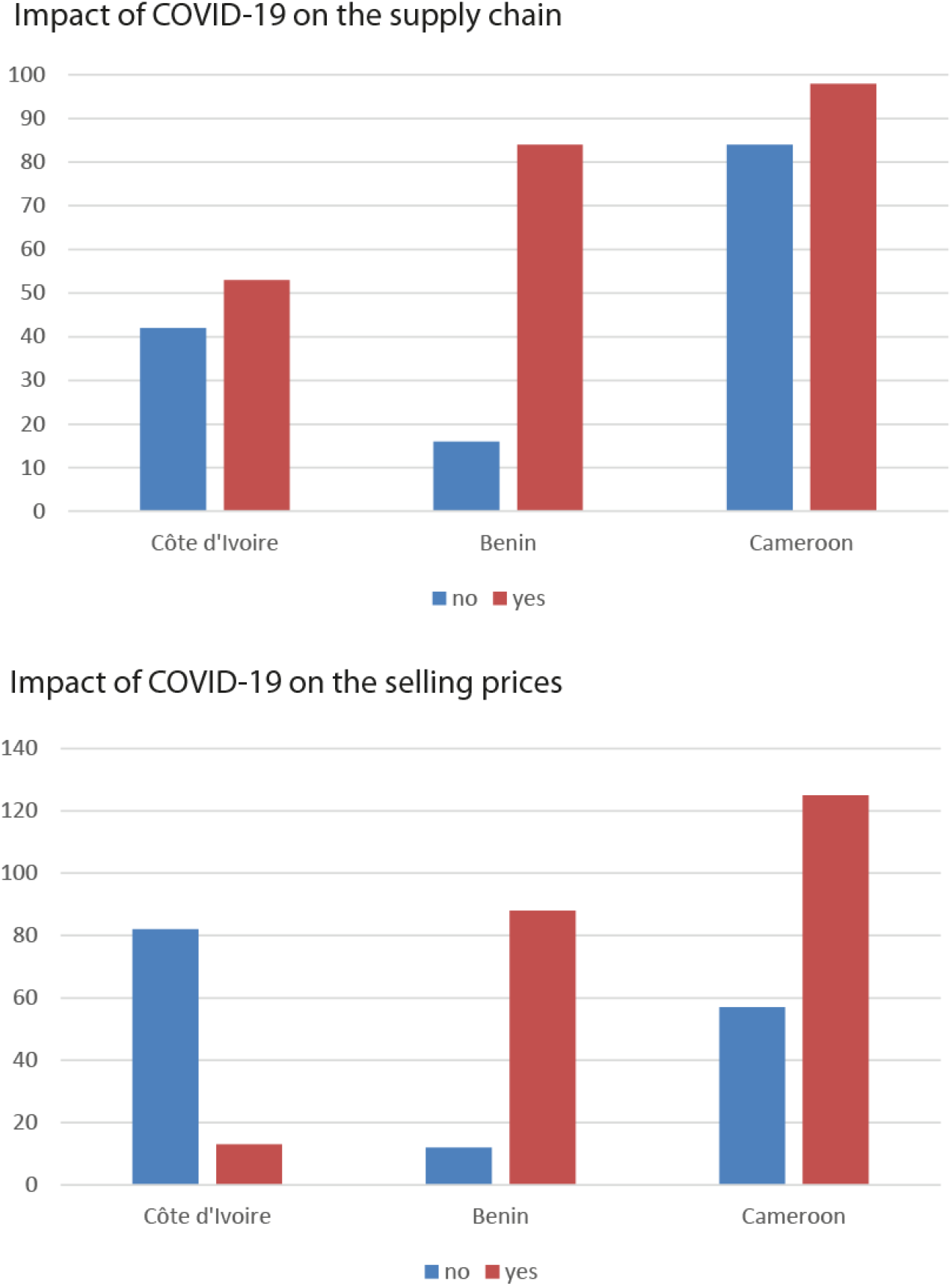
Perceived (negative) impacts of the COVID-19 pandemic on the bushmeat trade supply chain and selling prices among the vendors surveyed in Côte d’Ivoire, Benin and Cameroon.

**Appendix Figure 8.**
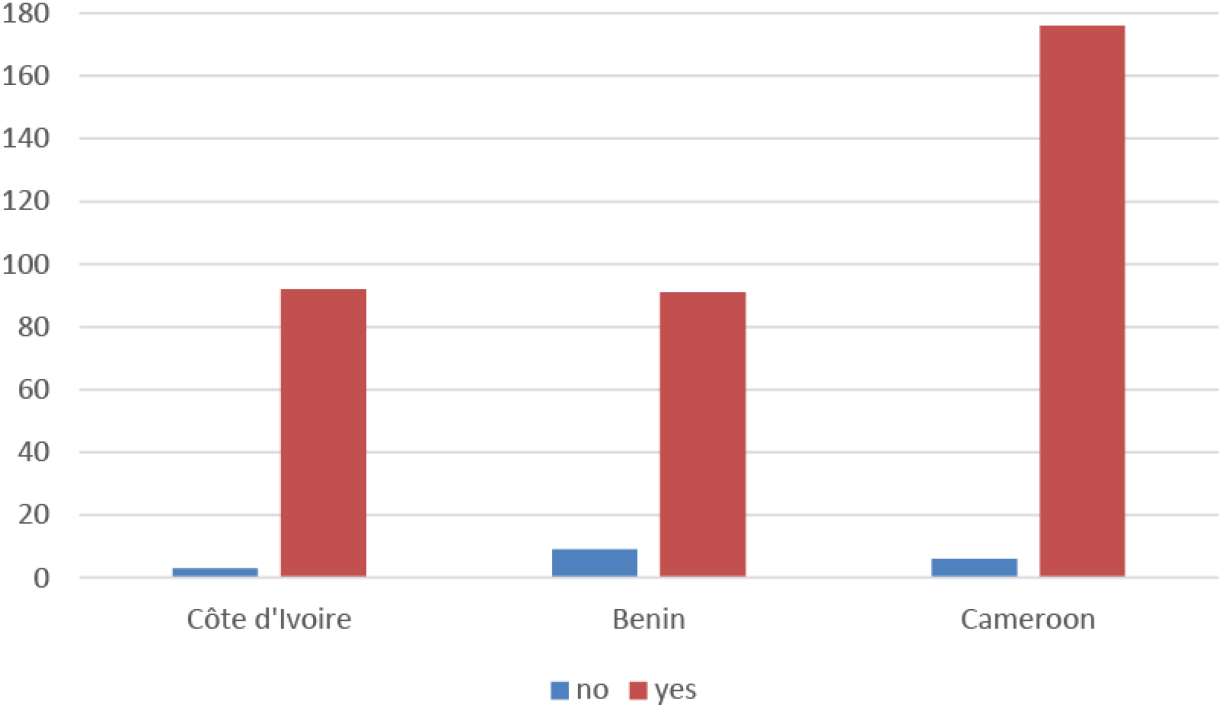
Number of bushmeat vendors reporting on their will to continue their activities despite the COVID-19 pandemic in Côte d’Ivoire, Benin and Cameroon.

**Appendix Figure 9.**
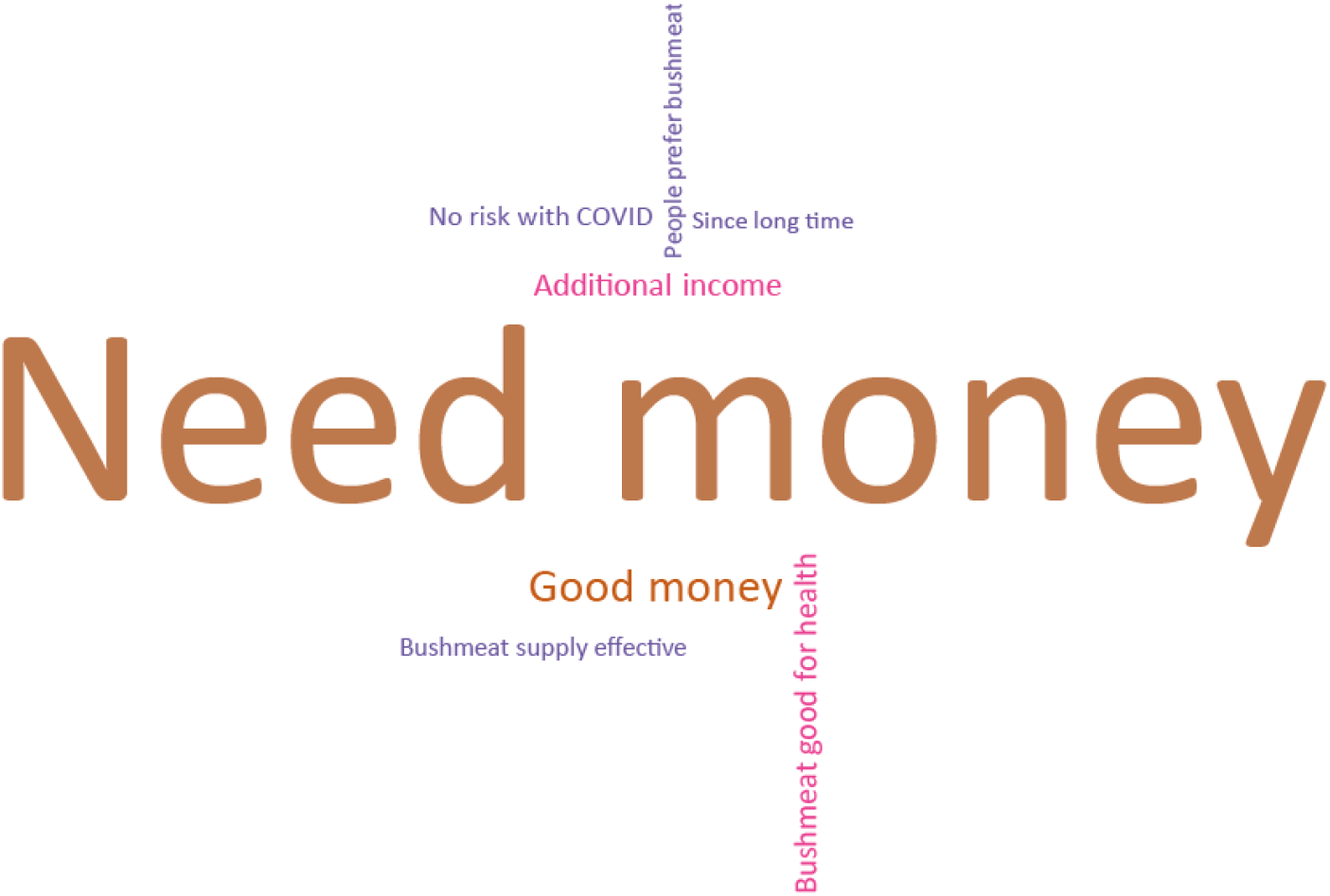
Word cloud representation of the reasons given by the bushmeat vendors to continue their activities despite the COVID-19 pandemic.

**Appendix Figure 10.**
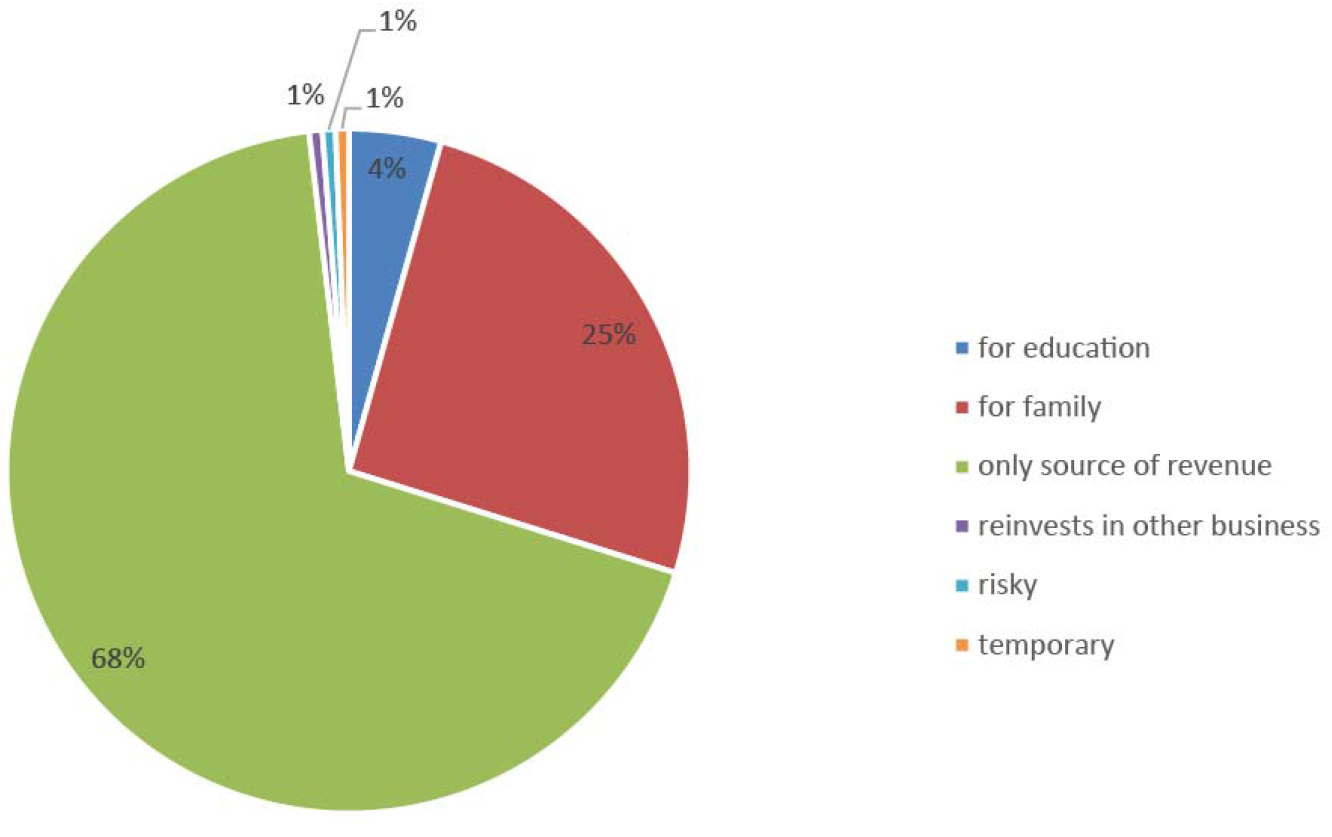
Justifications given by the bushmeat traders for being involved in their commercial activity.

**Appendix Figure 11.**
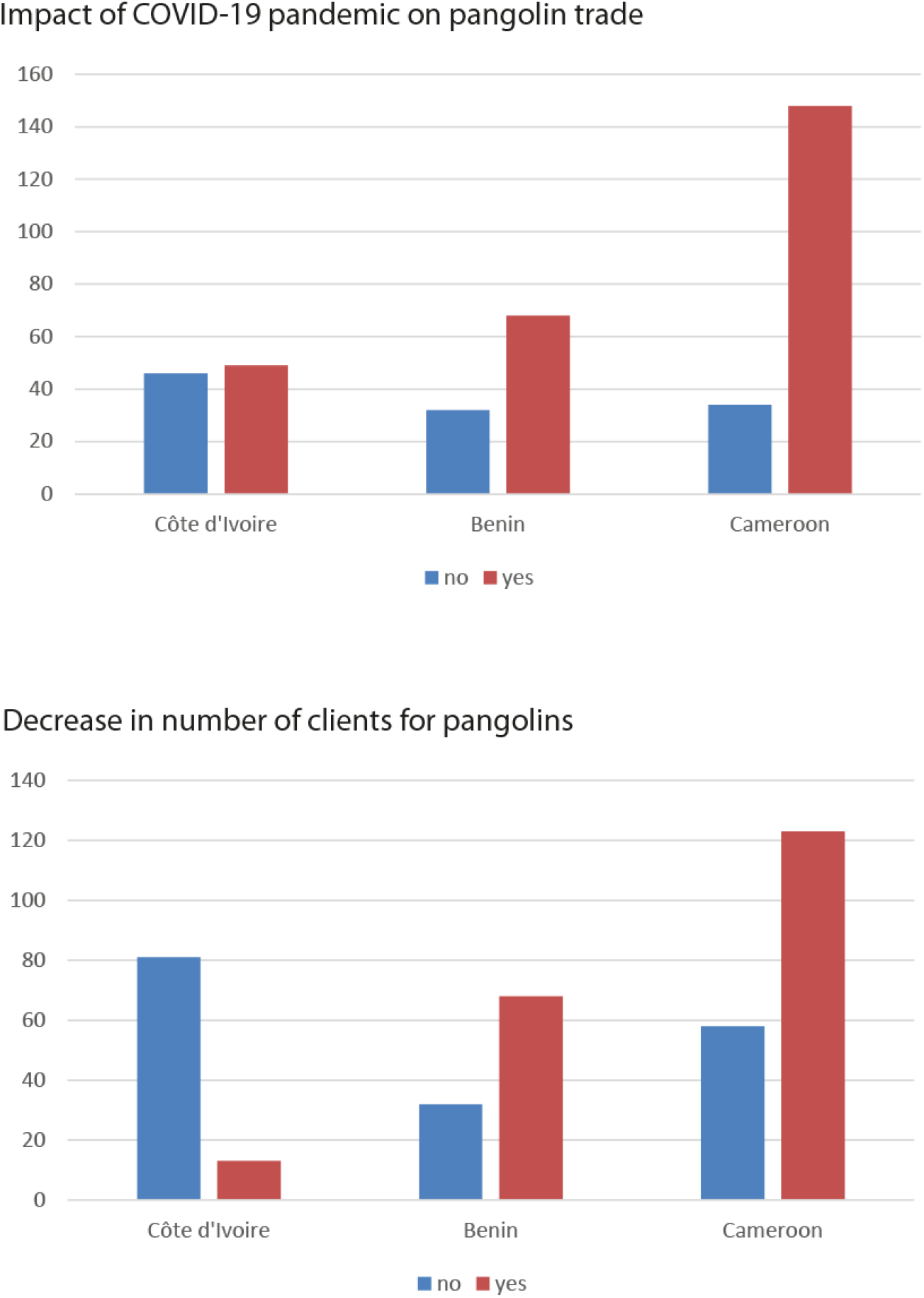
Perceived (negative) impacts of the COVID-19 pandemic on the pangolin trade dynamics among the vendors surveyed in Côte d’Ivoire, Benin and Cameroon.

**Appendix Figure 12.**
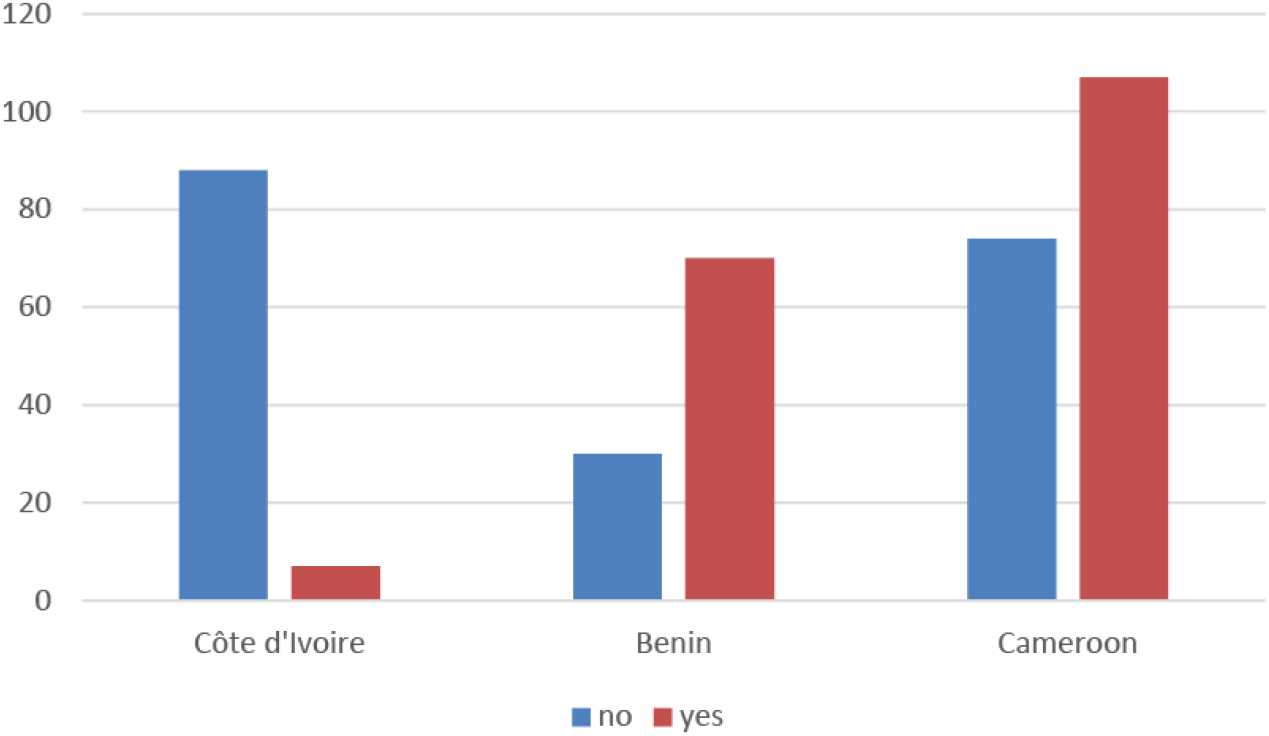
Perceived (negative) impact of the COVID-19 pandemic on the selling prices of pangolins among the vendors surveyed in Côte d’Ivoire, Benin and Cameroon.

**Appendix Figure 13.**
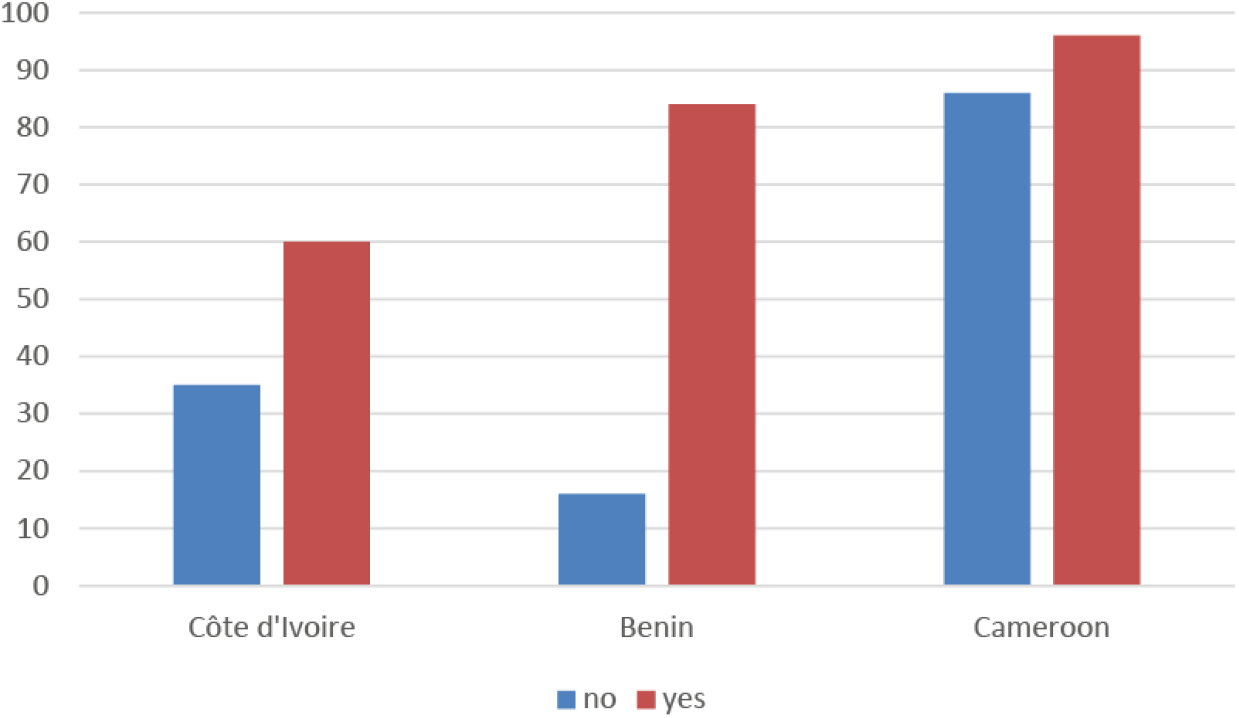
Number of bushmeat vendors reporting on whether state authorities took measures to control the market in Côte d’Ivoire, Benin and Cameroon.

**Appendix Figure 14.**
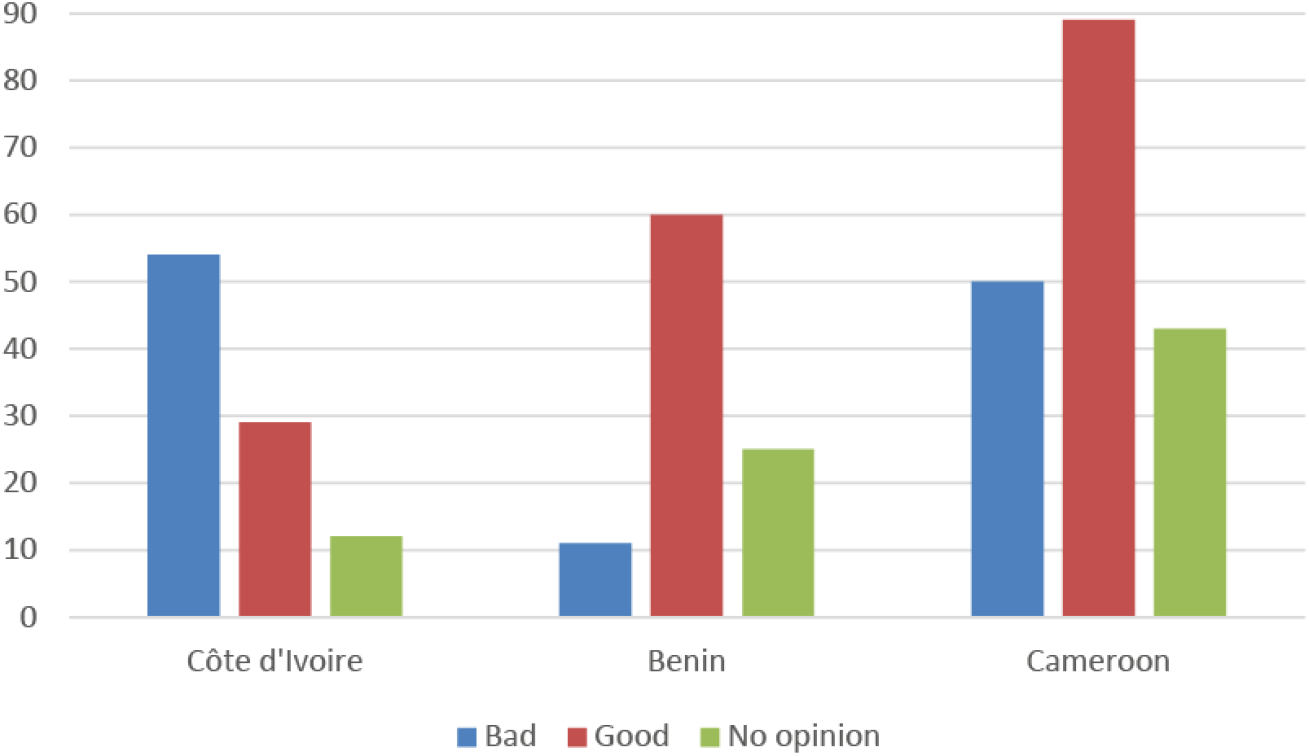
Perceptions on governmental measures against the COVID-19 pandemic among the bushmeat vendors in Côte d’Ivoire, Benin and Cameroon.

**Appendix Figure 15.**
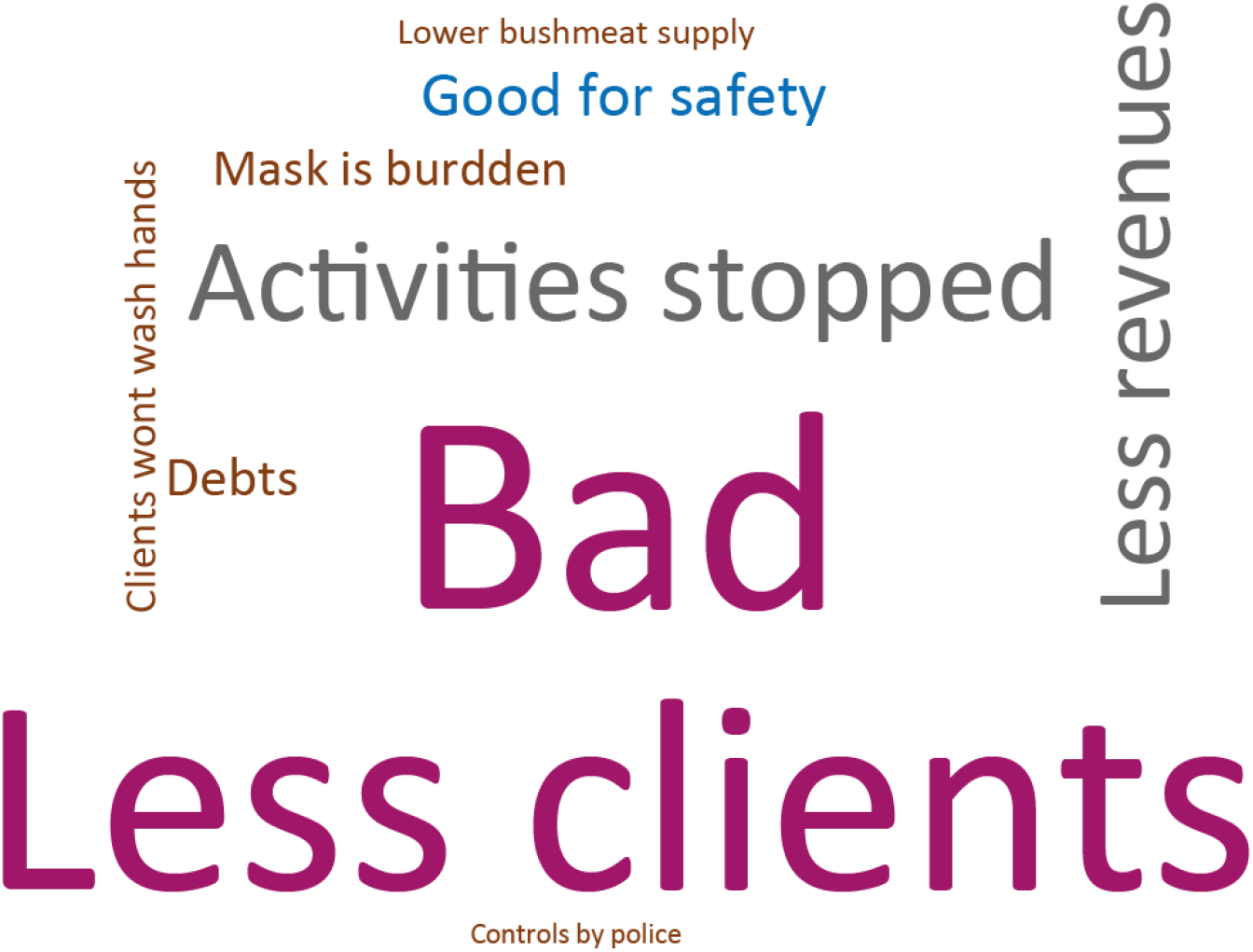
Word cloud representation of the perceptions on governmental measures against the COVID-19 pandemic among bushmeat vendors across three western African countries (Côte d’Ivoire, Benin and Cameroon). In blue, positive perceptions. In non-blue, negative perceptions. Word size is correlated to the number of times the answer was given. Example: Bad – 66 times; Good for safety – 10 times.

**Appendix Figure 16.**
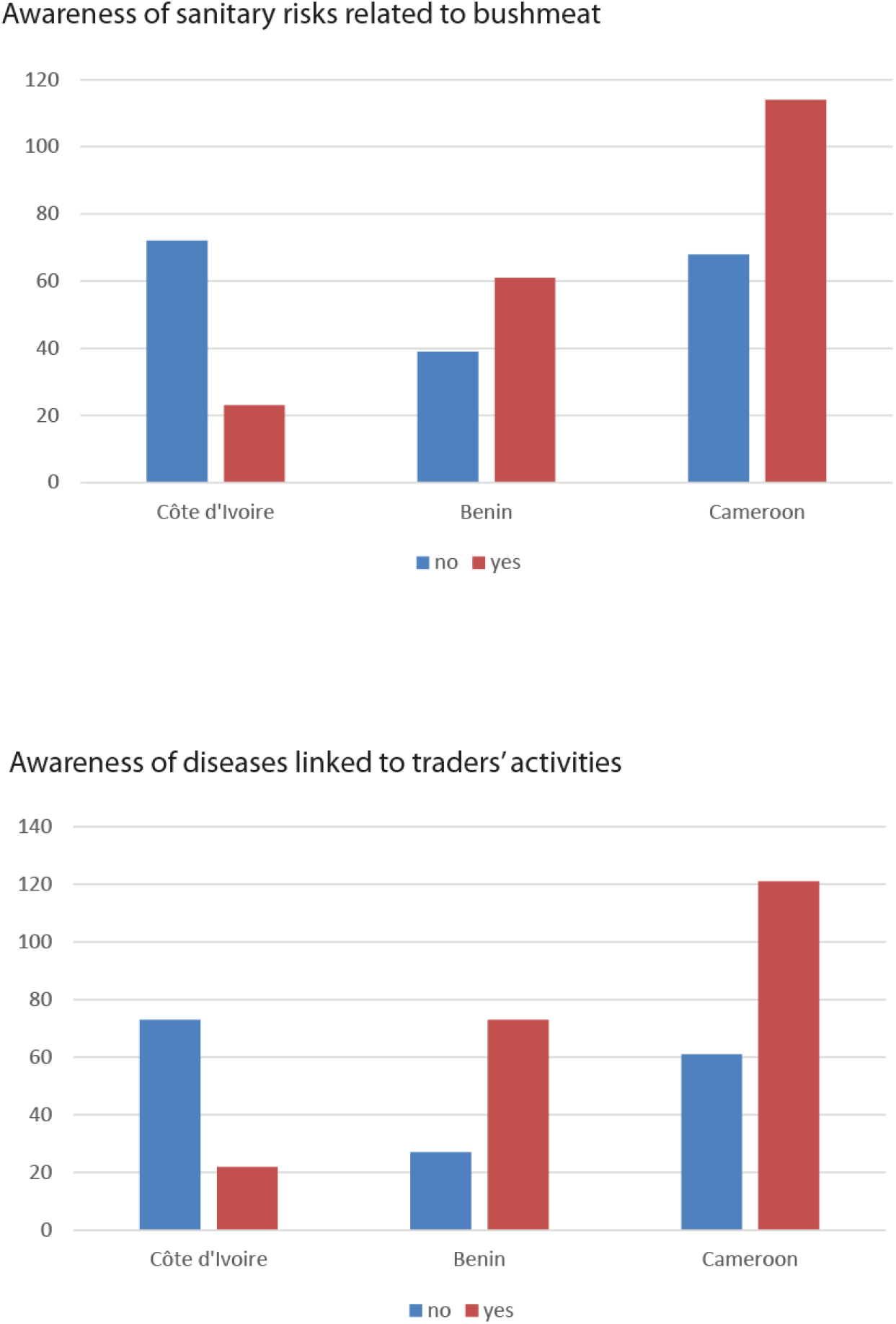
Health risk awareness among bushmeat vendors from Côte d’Ivoire, Benin and Cameroon.

**Appendix Figure 17.**
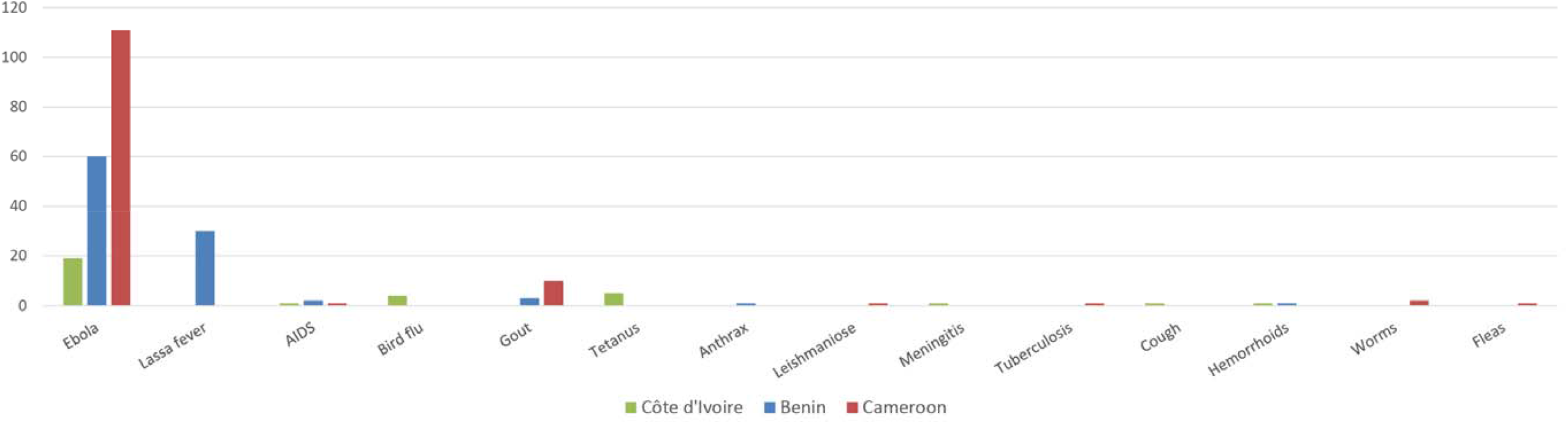
Diseases (COVID-19 excluded) transmitted by the bushmeat according to vendors from Côte d’Ivoire, Benin and Cameroon.

**Appendix Figure 18.**
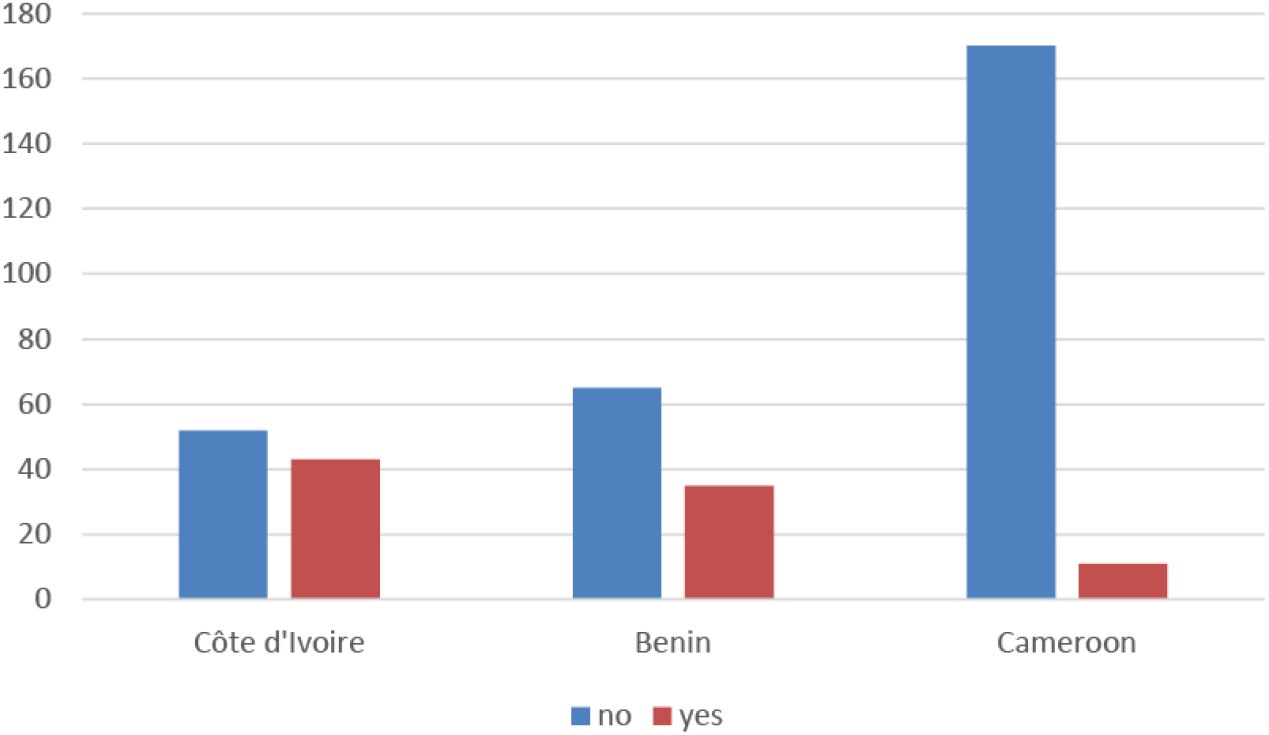
Number of bushmeat vendors reporting on being sick during their periods of activity in Côte d’Ivoire, Benin and Cameroon.

**Appendix Figure 19.**
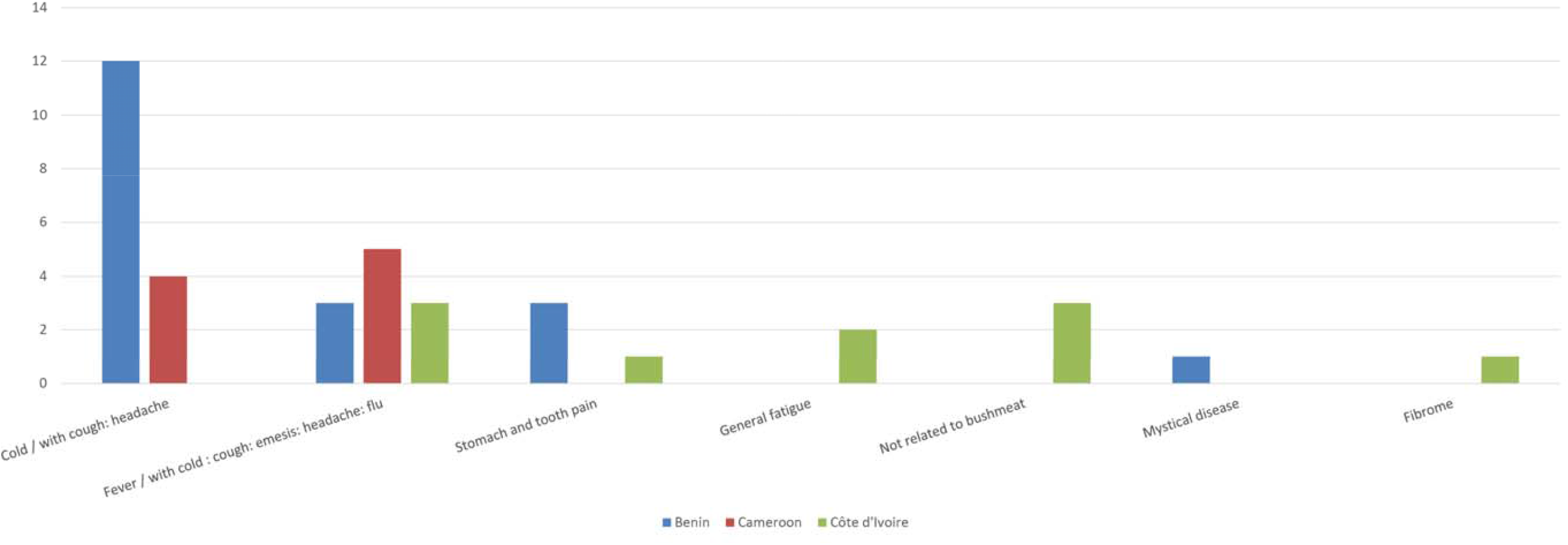
Sickness symptoms reported by bushmeat vendors sick during their periods of activity in Côte d’Ivoire, Benin and Cameroon.

**Appendix Figure 20.**
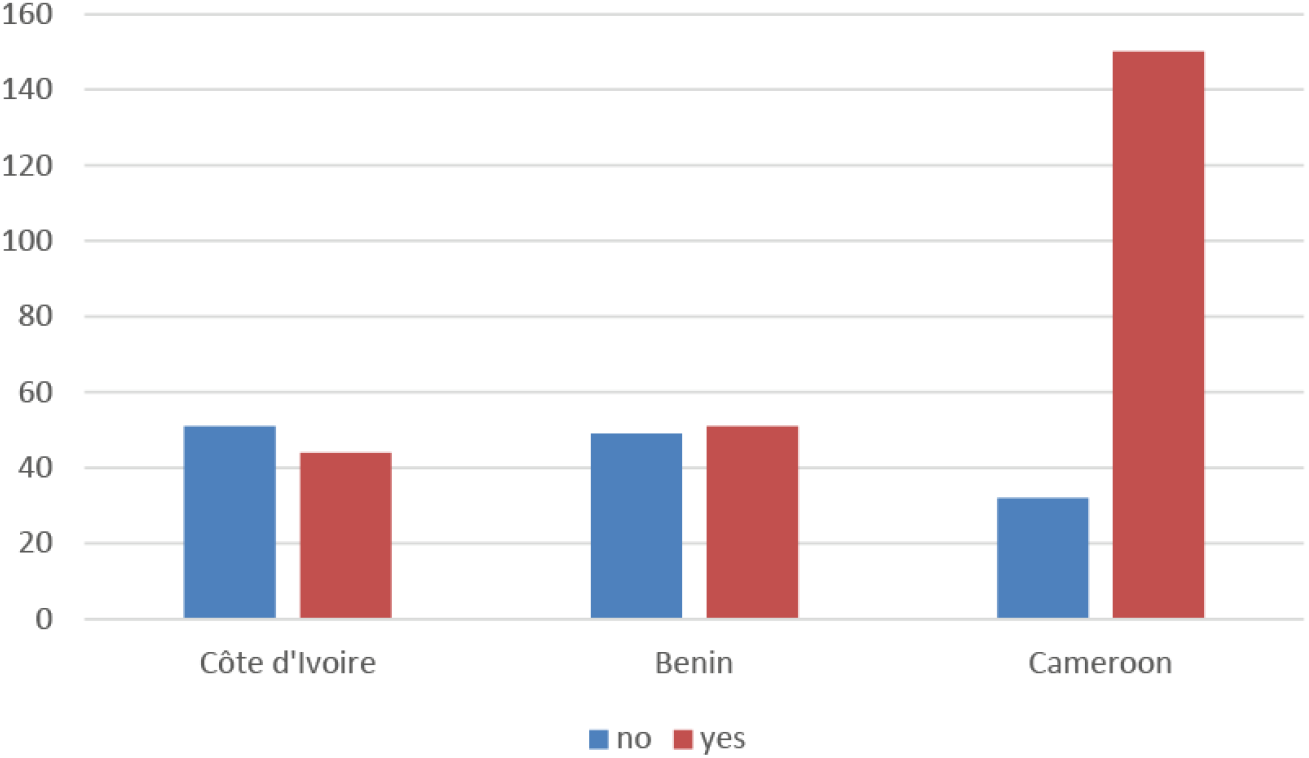
Number of bushmeat vendors reporting whether they were aware of the information that pangolins could be involved in the COVID-19 pandemic in Côte d’Ivoire, Benin and Cameroon.

**Appendix Figure 21.**
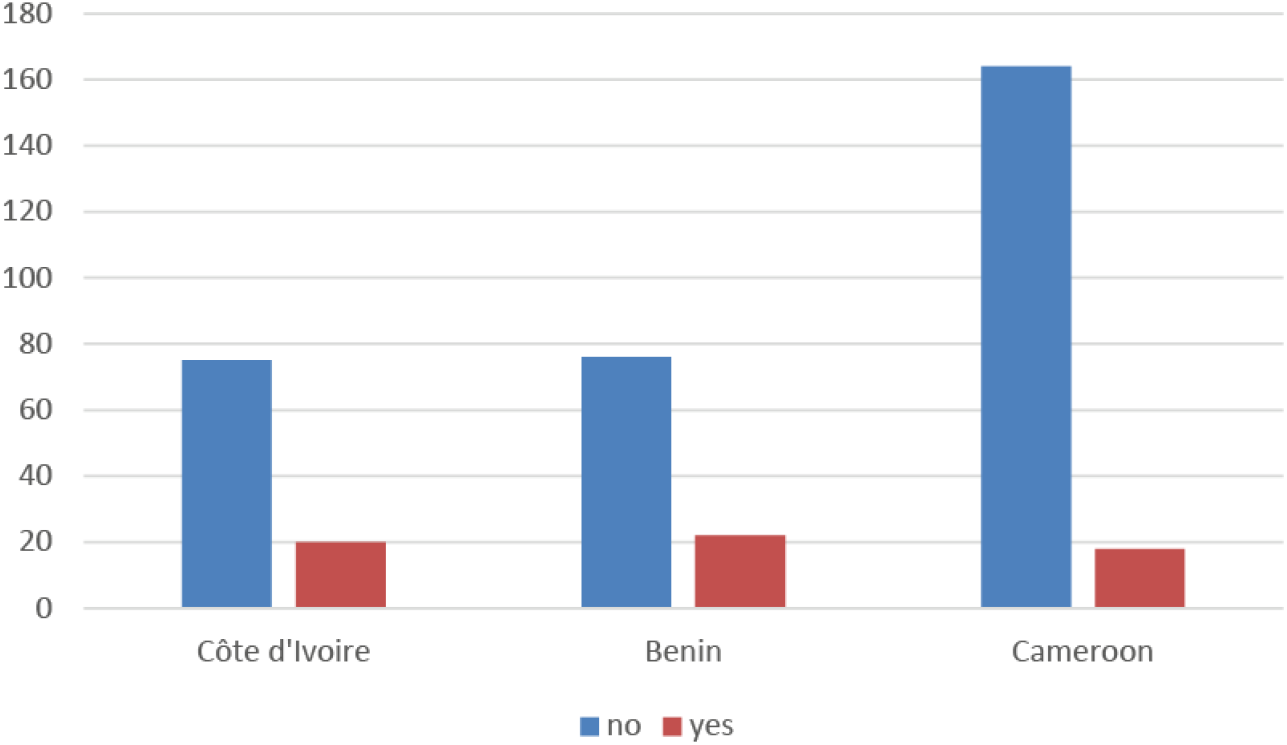
Number of bushmeat vendors reporting whether they believe that pangolins are involved in the COVID-19 pandemic in Côte d’Ivoire, Benin and Cameroon.

**Appendix Document 1**. Semi-structured questionnaire used to study the impact of the COVID-19 pandemic on the bushmeat trade in three western African countries (Côte d’Ivoire, Benin, Cameroon). The questionnaire was addressed to 377 vendors from 48 bushmeat trade sites.

Readers may contact the corresponding author for accessing the original French version of the questionnaire.

<u>A- Sociodemographic variables:</u>

1. Age:
2. Gender:
3. Ethnic group:
4. Education level:
5. Seniority in the trade: 6. Locality / market:

<u>B- Questionnaire:</u>

1. Did the COVID-19 pandemic have an impact on your activities? *☐Yes ☐ No*

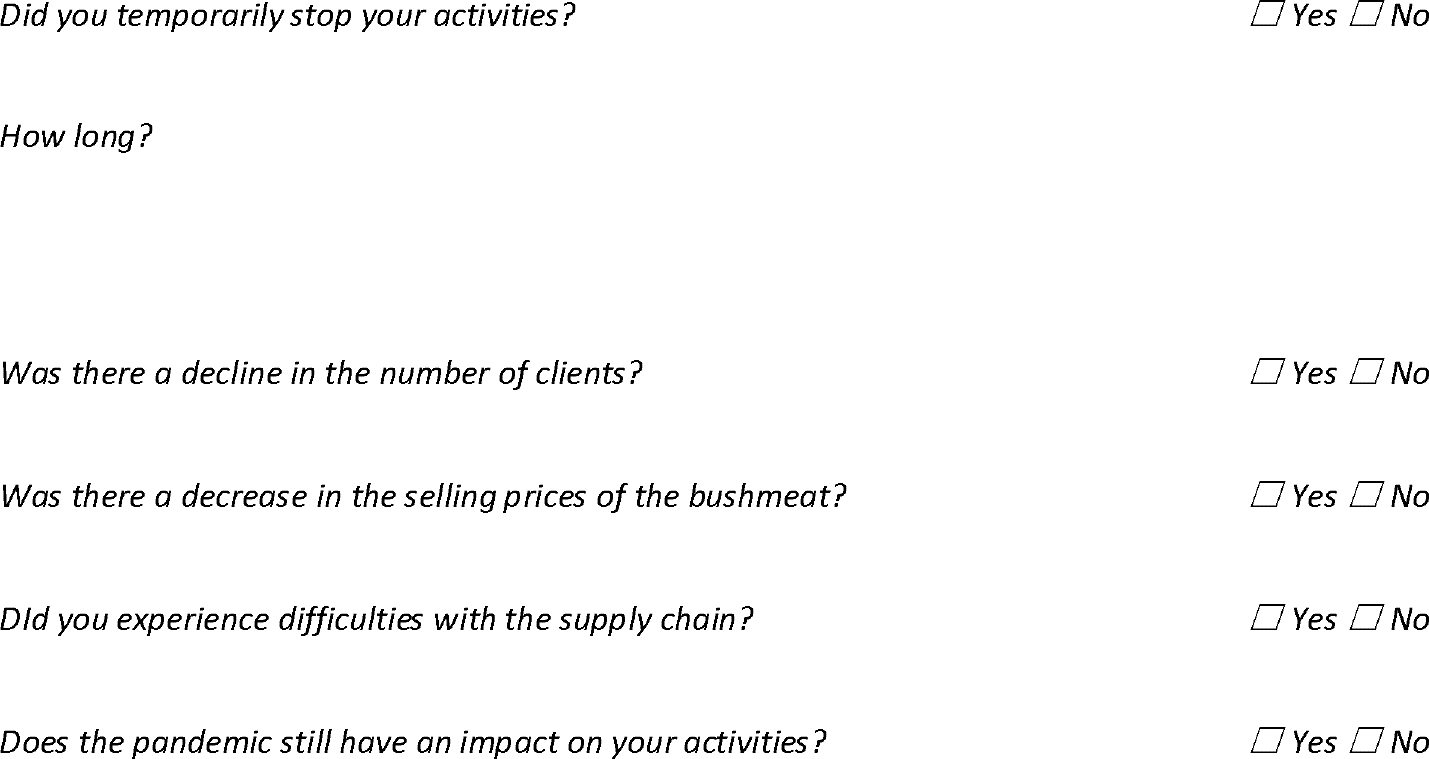 *\*Supplementary information*
2. Did you know that pangolins could be involved in the COVID-19 pandemic? *Yes ☐ No* *Source of information?*
3. Do you believe in this information ? *Yes ☐ No* *Why?*
4. Did the COVID-19 pandemic have an impact of the pangolin trade? *☐ Yes ☐ No*

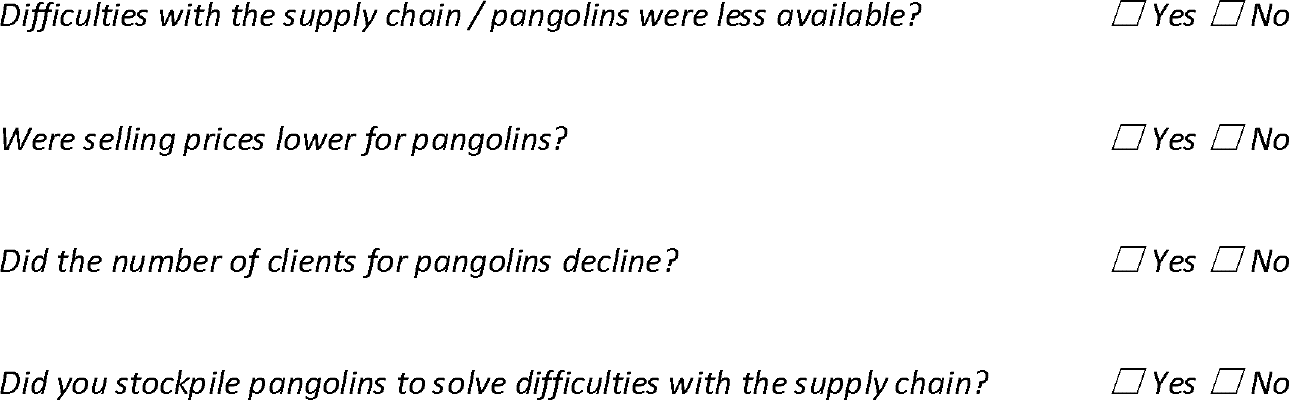 *\*Supplementary information*
5. Do you know that sanitary risks are linked to your activity? *☐ Yes ☐ No*

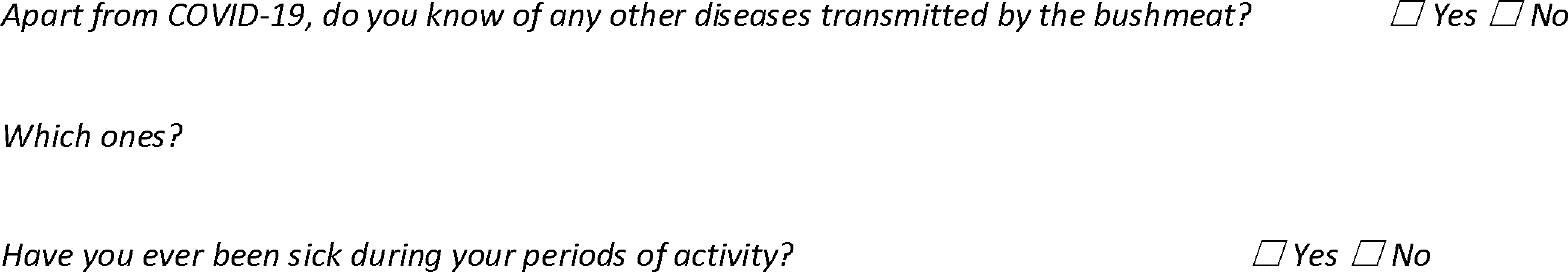 *Which were the symptoms / diseases?* *Supplementary information
6. Were there measures taken by the authorities to control the markets? *Yes ☐ No* *What do you think of the measures taken by the government to fight against the COVID-19?* *☐ Good ☐ Bad ☐ No opinion* *What were these measures?* *What was the impact of these measures on your activities?*
7. Despite the effects of the pandemic, do you wish to continue your activities? *Yes ☐ No* *Why?* *\*Supplementary information*

